# SurvivEHR: a competing risks, time-to-event foundation model for multiple long-term conditions from primary care electronic health records

**DOI:** 10.1101/2025.08.04.25332916

**Authors:** Charles Gadd, Krishna Gokhale, Aditya Acharya, Jennifer Cooper, Francesca Crowe, Leah Fitzsimmons, Thomas Jackson, Krishnarajah Nirantharakumar, Christopher Yau, The OPTIMAL collaborative

## Abstract

Multiple long-term conditions (MLTCs) or multimorbidity – the co-occurrence of multiple chronic conditions –presents a growing challenge for primary care. Current predictive models often target single outcomes and overlook the complexities of time-to-event risk in real-world, longitudinal health data. Here, we present SurvivEHR, a generative transformer-based foundation model trained on over 7.6 billion coded events from 23 million patients in UK primary care. SurvivEHR introduces a competing risk time-to-event pretraining objective that enables accurate forecasting of future diagnoses, investigations, medications, and mortality. We demonstrate that SurvivEHR achieves strong risk stratification performance, captures clinically meaningful trajectories, and outperforms benchmark survival models across multiple tasks. The model also transfers effectively to fine-tuned prognostic tasks, particularly in low-resource settings. By learning patient trajectories directly from routine health records, SurvivEHR offers a scalable and privacy-preserving approach for building generalisable clinical risk tools that address the complexity of MLTCs in primary care.

## Introduction

The study of Multiple Long-Term Conditions (MLTCs), also referred to as multimorbidity, is becoming increasingly important as the prevalence of individuals living with two or more chronic conditions continues to rise [1]. This shift is largely driven by an aging population and advances in medical care that have extended life expectancy, resulting in more people living longer with chronic diseases [2]. MLTCs are now the norm rather than the exception in many healthcare settings, particularly in high-income countries, and are associated with poorer health outcomes, reduced quality of life, increased healthcare costs, and higher rates of hospitalisation and mortality [3, 4]. Despite the scale of the issue, healthcare systems, research frameworks, and clinical guidelines often remain structured around single-disease models, which do not reflect the complex realities faced by patients with MLTCs [5].

In many health systems, primary care plays a central role in the management of MLTCs, serving as the first point of contact for most patients and providing continuous, coordinated care across multiple specialities. General practitioners (GPs) are uniquely positioned to understand the broader context of a patient’s health, including the interplay between physical, mental, and social factors. However, the growing complexity of care required by patients with MLTCs places significant strain on primary care services and exposes gaps in the evidence base that underpins clinical decision-making [6]. Effective management of MLTCs in primary care requires a shift away from disease-specific approaches toward more holistic, patient-centred models that consider the cumulative and interactive effects of multiple conditions [5].

Many current clinical risk prediction models that are in use remain focused on single conditions such as cardiovascular disease [7, 8], diabetes [9], or cancer [10]. While these models can be useful in targeting disease-specific interventions, they are not designed to account for the wider health status of patients and in fact often only accept input of the patient’s current characteristics. This may lead to missed opportunities for improved care and prevention [2] and there is a need to develop and implement risk prediction tools that better reflect the complexity of MLTCs, particularly in primary care where early identification and proactive management can be most effectively implemented.

Deep learning-based (DL) models for electronic health records (EHRs) [11] has enabled a new generation of EHR-based predictive models based on coded data to be constructed that overcome the limitations of traditional risk prediction models. Predictive models using cross-sectional (non-longitudinal) inputs such as Deephit [12] and DeSurv [13] can provide competing risk, time-to-event models to flexibly handle heterogeneous input features. However, models that specifically utilise longitudinal inputs (Table 1) are capable of learning from and using entire patient histories thus potentially accounting for complex MLTC backgrounds. Some models can also be considered to be *foundation models* learn general patterns from EHR data that be used as a basis for specific prediction tasks via transfer learning [14].

**Table 1.**
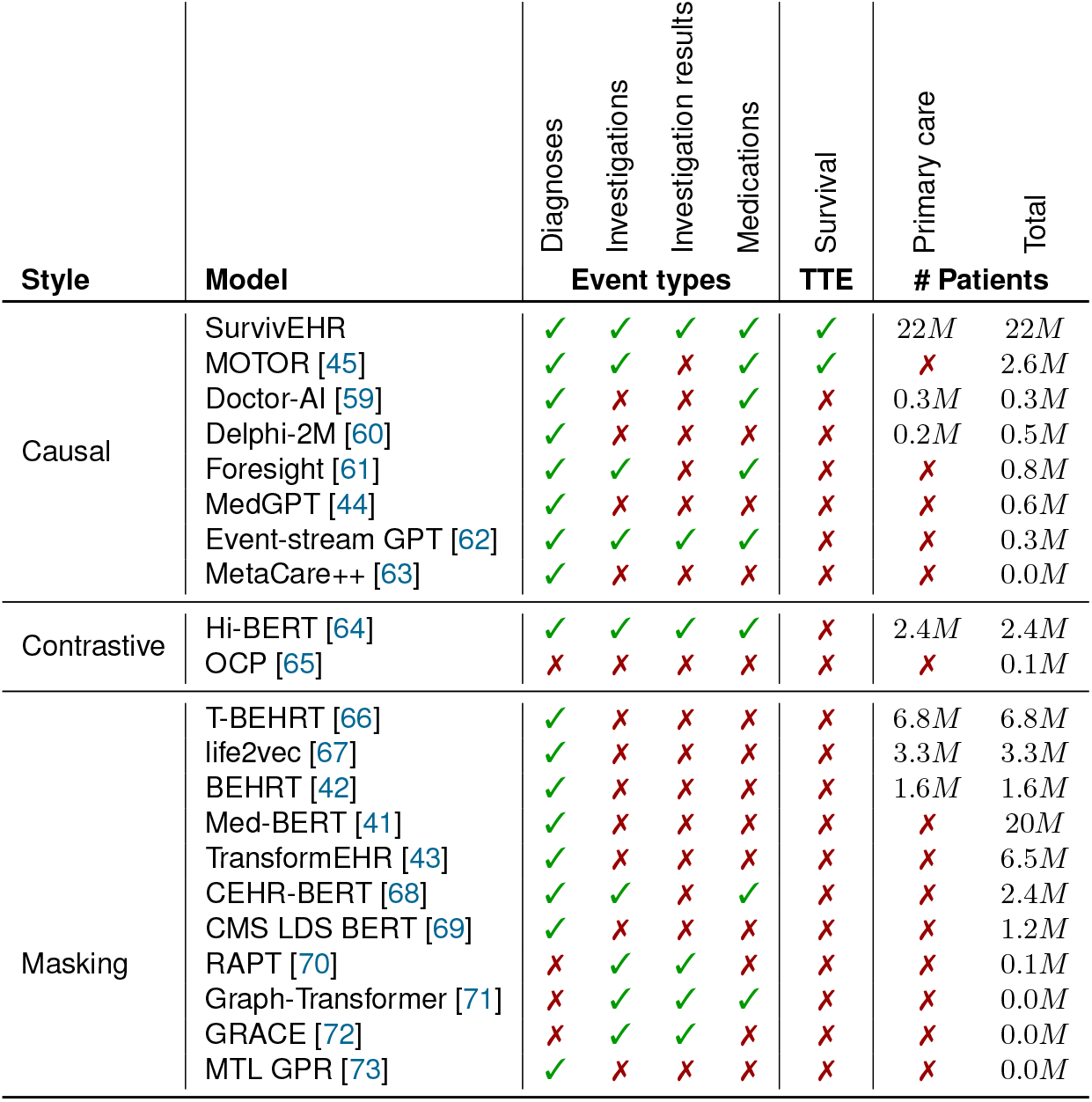
A summary of existing deep learning-based clinical risk prediction models. TTE (time-to-event) refers to learning the full distribution of the time-to-event, rather than ignoring time or making only a point prediction. Information not made available is left blank

| Style | Model | Diagnoses | Investigations | Investigation results | Medications | Survival | Primary care | Total |
| --- | --- | --- | --- | --- | --- | --- | --- | --- |
|  |  | Event types |  |  |  | TTE | # Patients |  |
| Causal | SurvivEHR | ✓ | ✓ | ✓ | ✓ | ✓ | 22M | 22M |
|  | MOTOR [45] | ✓ | ✓ | ✗ | ✓ | ✓ | ✗ | 2.6M |
|  | Doctor-AI [59] | ✓ | ✗ | ✗ | ✓ | ✗ | 0.3M | 0.3M |
|  | Delphi-2M [60] | ✓ | ✗ | ✗ | ✗ | ✗ | 0.2M | 0.5M |
|  | Foresight [61] | ✓ | ✓ | ✗ | ✓ | ✗ | ✗ | 0.8M |
|  | MedGPT [44] | ✓ | ✗ | ✗ | ✗ | ✗ | ✗ | 0.6M |
|  | Event-stream GPT [62] | ✓ | ✓ | ✓ | ✓ | ✗ | ✗ | 0.3M |
|  | MetaCare++ [63] | ✓ | ✗ | ✗ | ✗ | ✗ | ✗ | 0.0M |
| Contrastive | Hi-BERT [64] | ✓ | ✓ | ✓ | ✓ | ✗ | 2.4M | 2.4M |
|  | OCP [65] | ✗ | ✗ | ✗ | ✗ | ✗ | ✗ | 0.1M |
| Masking | T-BEHRT [66] | ✓ | ✗ | ✗ | ✗ | ✗ | 6.8M | 6.8M |
|  | life2vec [67] | ✓ | ✗ | ✗ | ✗ | ✗ | 3.3M | 3.3M |
|  | BEHRT [42] | ✓ | ✗ | ✗ | ✗ | ✗ | 1.6M | 1.6M |
|  | Med-BERT [41] | ✓ | ✗ | ✗ | ✗ | ✗ | ✗ | 20M |
|  | TransformEHR [43] | ✓ | ✗ | ✗ | ✗ | ✗ | ✗ | 6.5M |
|  | CEHR-BERT [68] | ✓ | ✓ | ✗ | ✓ | ✗ | ✗ | 2.4M |
|  | CMS LDS BERT [69] | ✓ | ✗ | ✗ | ✗ | ✗ | ✗ | 1.2M |
|  | RAPT [70] | ✗ | ✓ | ✓ | ✗ | ✗ | ✗ | 0.1M |
|  | Graph-Transformer [71] | ✗ | ✓ | ✓ | ✓ | ✗ | ✗ | 0.0M |
|  | GRACE [72] | ✗ | ✓ | ✓ | ✗ | ✗ | ✗ | 0.0M |
|  | MTL GPR [73] | ✓ | ✗ | ✗ | ✗ | ✗ | ✗ | 0.0M |

However, despite the plethora of DL prediction models, limitations on data availability remain a challenge and has had unintentional consequences on the type of models developed. At the time of writing, we are only aware of a small number of DL models have been developed using primary care data^1^ and these predominantly originate from European institutions that make use of European data sources such as the UK Clinical Practice Research Datalink (CPRD) [15], Whole Systems Integrated Care (WSIC) Northwest London [16], UK Biobank [17]), the Finnish Registry [18], The Information System for Research in Primary Care (SIDIAP) in Spain [19, 20] and the Danish Health Registry data [21]. This is in contrast to the much wider family of DL models which have been developed using secondary care datasets including MIMIC-IV [22], US Veteran’s Health Administration data [23] as well as data from individual hospital (networks) and proprietary medical insurance claims (e.g. Truven Health MarketScan and Partners For Kids) databases.

Secondary care involves specialised treatment for more complex or severe conditions, typically following a referral from a primary care provider. The nature of clinical workflows and the resulting data structures differ substantially between these settings, leading to distinct considerations in the development of deep learning (DL) models. For example, consider acute kidney injury (AKI). In secondary care, patients may be monitored for AKI following events such as emergency hospital admission, major infections, or complications after surgery. The condition is more likely to be acute, rapidly progressing, and associated with significant morbidity. Patients often undergo frequent blood tests, sometimes multiple times per day, and may be managed in high-dependency environments such as intensive care units (ICUs) [24]. In contrast, AKI in primary care is more often community-acquired, typically at an earlier stage and may be less severe. It can result from infections, dehydration, or drug-related nephrotoxicity (e.g., from combinations like ACE inhibitors and NSAIDs). Detection often depends on identifying trends in serum creatinine from blood tests that may be spaced weeks or months apart, and urine output is rarely available [25, 26]. In general, secondary care data typically cover shorter episodes of care with high-resolution, densely collected clinical information, whereas primary care data offer lower temporal granularity but broader longitudinal coverage, often spanning decades of a patient’s health history.

Motivated by the lack of models specifically targeting primary care and working with our patient advisory group, we propose a novel foundation model for time-to-event forecasting using primary care-based EHR inputs focusing on multiple long-term conditions. Based on the Generative Pre-trained Transformer (GPT) [27] architecture - named SurvivEHR, we trained on 7.6 billion patient records from 23 million patients the UK Clinical Practice Research Datalink (CPRD) [15] primary care record database, SurvivEHR is capable of forecasting primary care records, providing personalised forecasts for risk of future chronic disease diagnoses, measurements, tests, medications and death. We demonstrate that SurvivEHR supports strong transfer learning, and can be used as a foundation model for clinical prediction modelling on a number of usage examples.

## Results

### Model overview

We introduce **SurvivEHR**, a decoder-only transformer-based foundation model designed for next-event prediction with time-to-event estimation in longitudinal primary care electronic health records (EHRs). The model is trained in a self-supervised generative fashion, learning to predict both the type and timing of the next clinical event, thereby enabling calibrated temporal risk stratification across multiple dependent outcome types (competing risks).

The input into SurvivEHR is a patient’s longitudinal history of clinical events – including diagnoses, prescriptions, and measurements – as a sequence of tokens (Figure 1). Each token is embedded based on the event type, any associated numerical value (e.g. test result) and its timestamp relative to the patient’s age (Figure 1B). This sequence is processed via a *multi-head attention transformer* producing a latent representation of the current patient state (Supplementary Figure S1). To predict the time to next event, SurvivEHR integrates a neural-based competing risks framework [13] which yields calibrated Cumulative Incidence Functions (CIFs) for each potential outcome, representing the risk that each event occurs before a certain time.

**Figure 1:**
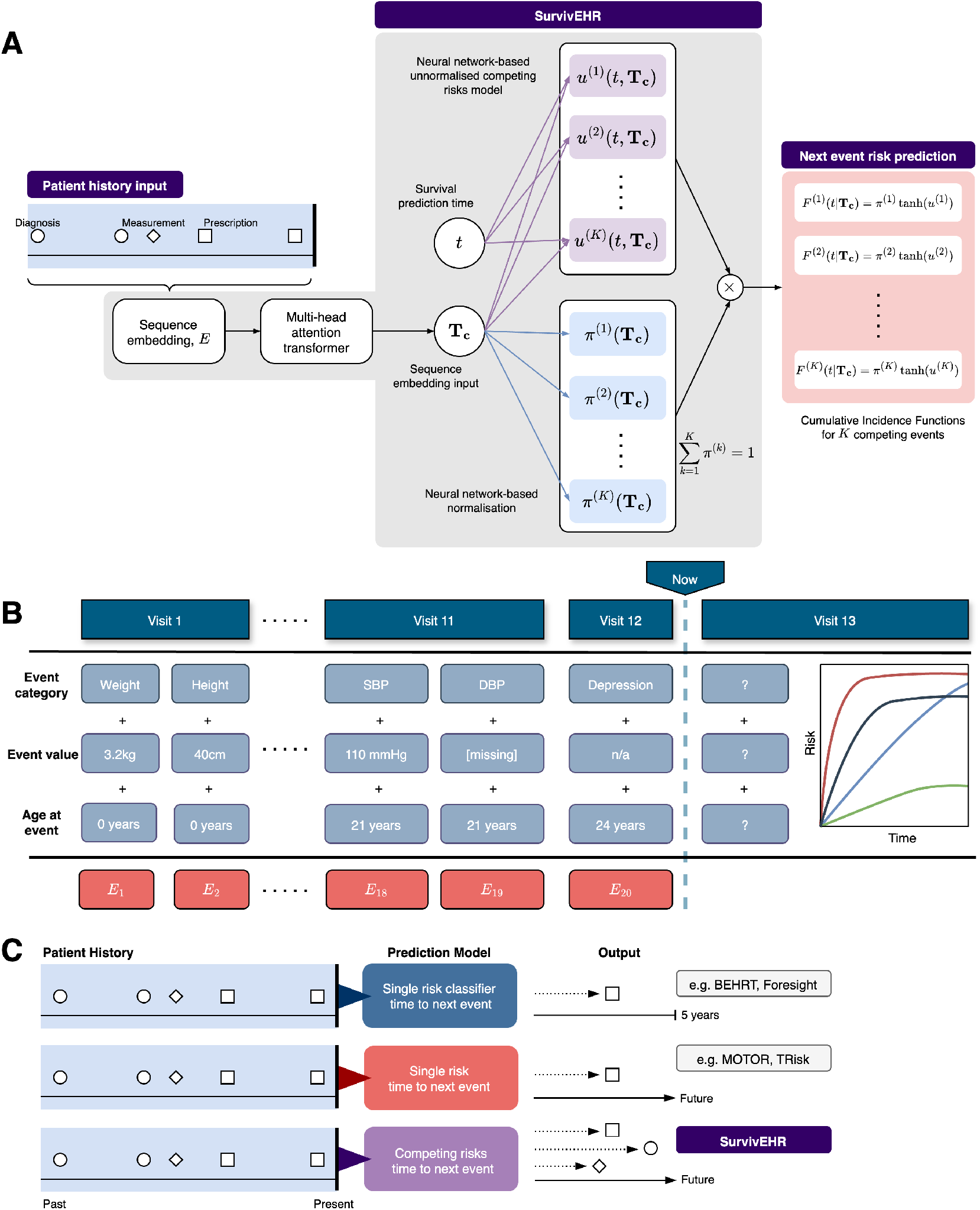
SurvivEHR. **A** Schematic overview of SurvivEHR model architecture and implementation. Sequence embedding of tokenised patient histories are fed to a multi-head attention transformer which is used within a neural network-based competing risk next-event prediction model. **B** Joint embeddings are created encoding event type, values and times to produce time-to-event competing risk and value prediction functions. **C** Competing risk time-to-event modelling distinguishes SurvivEHR from other related models that focus on discrete classification or single-risk modelling.

This distinguishes SurvivEHR from other DL EHR models in that its design is specifically catered to primary care and operates in a competing risk, time-to-event framework (Table 1). The approach is necessary since patterns of MLTCs in primary care are complex and diverse health events can occur over time scales across a person’s lifetime [28]. This makes the prediction problems different from that of predicting a single outcome risk, fixed period hospital readmission or disease recurrence and other problems which are well tackled by many existing DL EHR models.

### Data Overview

The data processing pipeline is schematically depicted in Figure 2A. We used the Clinical Practice Research Datalink (CPRD), a large longitudinal primary care database covering over 2,000 UK general practices using EMIS Web^®^. Around 10 million currently registered patients and 30 million historical records are included, broadly representative of the UK population. CPRD access was granted via the “OPTIMising therapies, disease trajectories, and AI assisted clinical management for patients Living with complex multimorbidity” (OPTIMAL) study.

**Figure 2:**
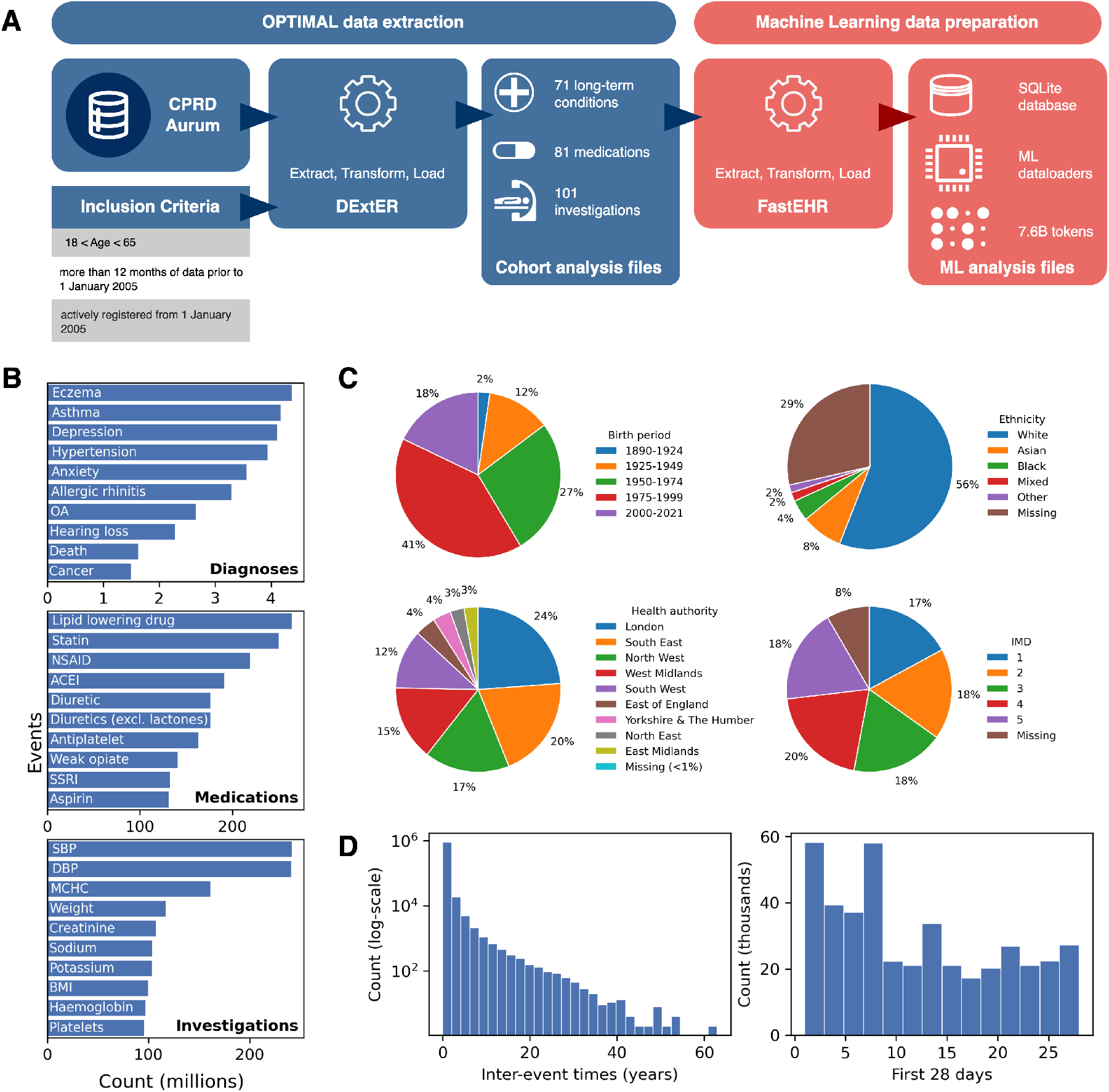
Overview of OPTIMAL cohort data preparation and content. **A** Data is extracted from CPRD Aurum using DExtER to produce a series of cohort analysis files. FastEHR is used to prepare the data for machine learning analysis. **B** Counts of the occurrence of different diagnoses, prescriptions and investigations. **C** Demographics summary of the individuals contained in the OPTIMAL cohort. **D** Distribution of inter-event times (left) all inter-event times (right) inter-event times within the first 28 days.

Data were extracted using DExtER [29]. Diagnoses, medications, and investigations were recorded using Read V2, SNOMED-CT, and EMIS codes. Pre-processing was conducted using FastEHR, a Python toolkit for scalable, memory-efficient EHR pipelines, enabling tokenisation, filtering, and leakage control. Post-processing, we retained 51M diagnoses, 4B medications, and 3.5B test records, 3.4B with numerical values. We applied a 90-5-5 split by practice site to avoid leakage, resulting in 23.6M training, 1.4M validation, and 1.5M test patients.

This work focuses on multiple long-term conditions and we identified 74 long-term conditions, 81 medication classes (based on DM&D Prodcodes), and 108 test types including blood tests and physiological measurements. The most prevalent items are shown in Figure 2B. Selection was informed by a Delphi study identifying 59 key conditions for multimorbidity research [30], and refined through expert and patient advisory input [31]. We included English patients with at least 12 months of acceptable data before 1 January 2005, yielding 26.5 million patients from 1,478 practices. The data covered a wide age, gender, ethnicity, geographical and deprivation (IMD) profile (Figure 2C). As discussed previously, inter-event intervals spanned a range from days to years which is consistent with lifelong patient history recording in primary care data (Figure 2D).

### Next event prediction performance

The evaluation of the pre-training performance of a foundation model is a critical step in understanding its capabilities and limitations. Proper evaluation metrics allow us to examine how well the pre-trained model aligns with downstream application requirements, and provides insights into its utility as a transferable learning framework. We evaluated the performance of SurvivEHR on its core task of risk-stratification for the next event given a patient history, introducing a new performance metric - the Inter-Event Concordance (IEC) metric - to assess risk discrimination in self-supervised survival modelling (see Methods).

We compare our pre-training strategy with two alternatives. Firstly, we consider an entirely prevalence based risk policy in which we prognosticate with a risk proportional to an event’s prevalence in the population. Secondly, we consider an alternative foundation model built upon the same backbone GPT architecture, but using a binary classification, cross-entropy pre-training objective. For this approach we assign the risk to be proportional to the classification probability.

SurvivEHR demonstrated strong self-supervised predictive performance compared to the baselines (Figure 3A, Supplementary Figure S2). We obtain a marginal IEC of 0.994, meaning that on average the true next event was among the top (1-0.994) × 263 ≈ 1.6 predicted events, demonstrating extremely strong overall predictive performance. Alternatively, following the baseline strategy of diagnosing by prevalence led to a score of 0.864, meaning that on average the true next event was among the top 36 most prevalent events. Finally, using the binary classification, cross-entropy training objective led to an IEC score of 0.699, meaning that on average the true next event was among the top 80 predicted illustrating that discretisation of a time-to-event problem can be detrimental.

**Figure 3:**
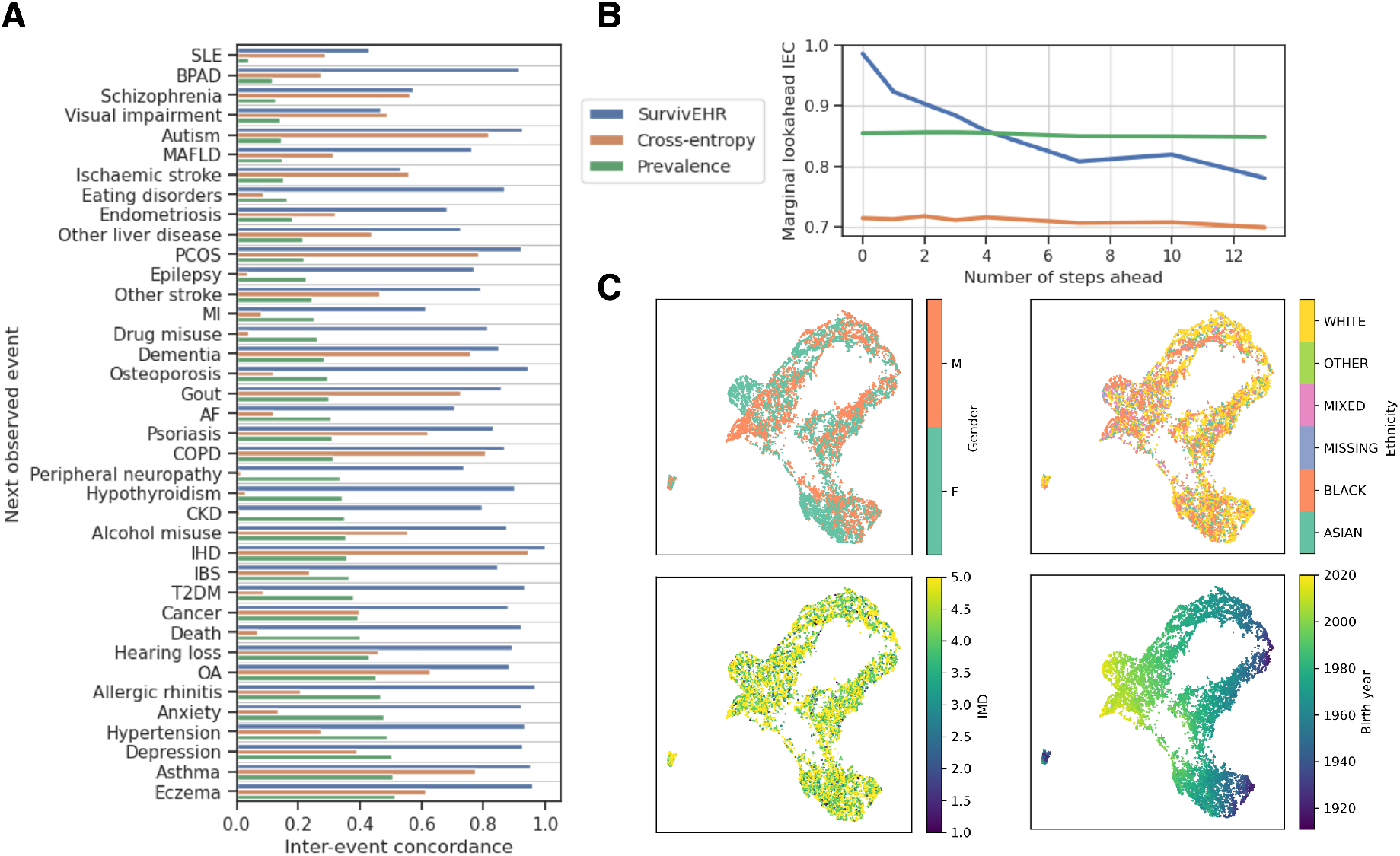
Evaluation of SurvivEHR on the pre-training dataset. **A** Each row gives the inter-event concordance score, which quantifies the model’s ability to correctly rank next-event risk within patient timelines. **B** Marginal inter-event concordance for predicting events increasingly into the future. As we predict further into the future we observe a reduction in the performance of foundation models. **C** UMAP representations for sex, ethnicity, index of multiple derivation (IMD), and age.

### Multi-step forecasting

During pre-training, SurvivEHR was tasked with predicting the next event. However, particularly when used for prognostic forecasting, we may be interested in the prediction of arbitrary future outcomes, rather than the immediate subsequent event. We tested the capacity of SurvivEHR to perform multi-step predictive forecasting in the pre-training test cohort. In Figure 3B, we used our marginal IEC score to evaluate how well SurvivEHR was able to stratify competing risks over an increasing time horizon, comparing this to the previous baseline approaches. Given a patient’s history, we used SurvivEHR to predict the first, second, third and beyond future events and observed that a clear degradation in performance occurs such that after four steps it is no longer able to outperform the prevalence-based baseline. This indicates a distributional shift between the pre-training (next step) and evaluation (four steps forward) tasks. Since SurvivEHR is built upon a modified GPT structure, it shares a conceptual parallel with challenges seen in typical LLMs in doing multi-step forecasting or long-horizon reasoning which have prompted recent developments in remedies such as chain-of-thought reasoning.

### Representation Learning and demographic bias

We next investigated, by visualisation, the 384-dimensional latent space used in SurvivEHR via UMAP [32] (Figure 3C, Supplementary Figure S3-4) to identify any clear emergent structures with respect to demographic variables. Visualisation yielded no obvious patterns with respect to male/female gender, ethnicity or index of multiple deprivation (IMD) indicating that predictions would not be heavily bias by these factors. The only clear associations were sex and birth year, as both will be significant predictive risk factors in most health conditions. We note that the lack of distinctive visualisable sub-structures likely reflects the heterogeneous and high complexity nature of the considerable proportion of the UK population in our CPRD data.

### SurvivEHR captures known clinical associations

We further explored if SurvivEHR is learning predictive patterns from the primary care records by identifying the clinical significance of the next event forecasts made by SurvivEHR. We use a recursive strategy to forecast a short sequence of future clinical trajectories and analyse the top marginal occurrence rates and the pairwise co-occurrence matrix that captures how often events arise together. Using the pre-training cohort, we present SurvivEHR with each held-out patient’s complete history and ask it to predict the next *three* clinical events.^2^ For every pair of events we compute the one-step transition probability from one event to the next event.

Figure 4A (Supplementary Figure S5) shows counts and conditional transition probabilities from SurvivEHR for an illustrative subset of diagnoses. Here there are strong conditional transitions for well-known clinical relationships such as Depression → Anxiety which is frequently observed in chronic mental health progression [33], Type 2 Diabetes (T2DM) → Osteoarthritis (OA) (obesity and systemic inflammation) [34] and Osteoarthritis (OA) → Myocardial infarction (MI) which is well-characterised epidemiological association [35].

**Figure 4:**
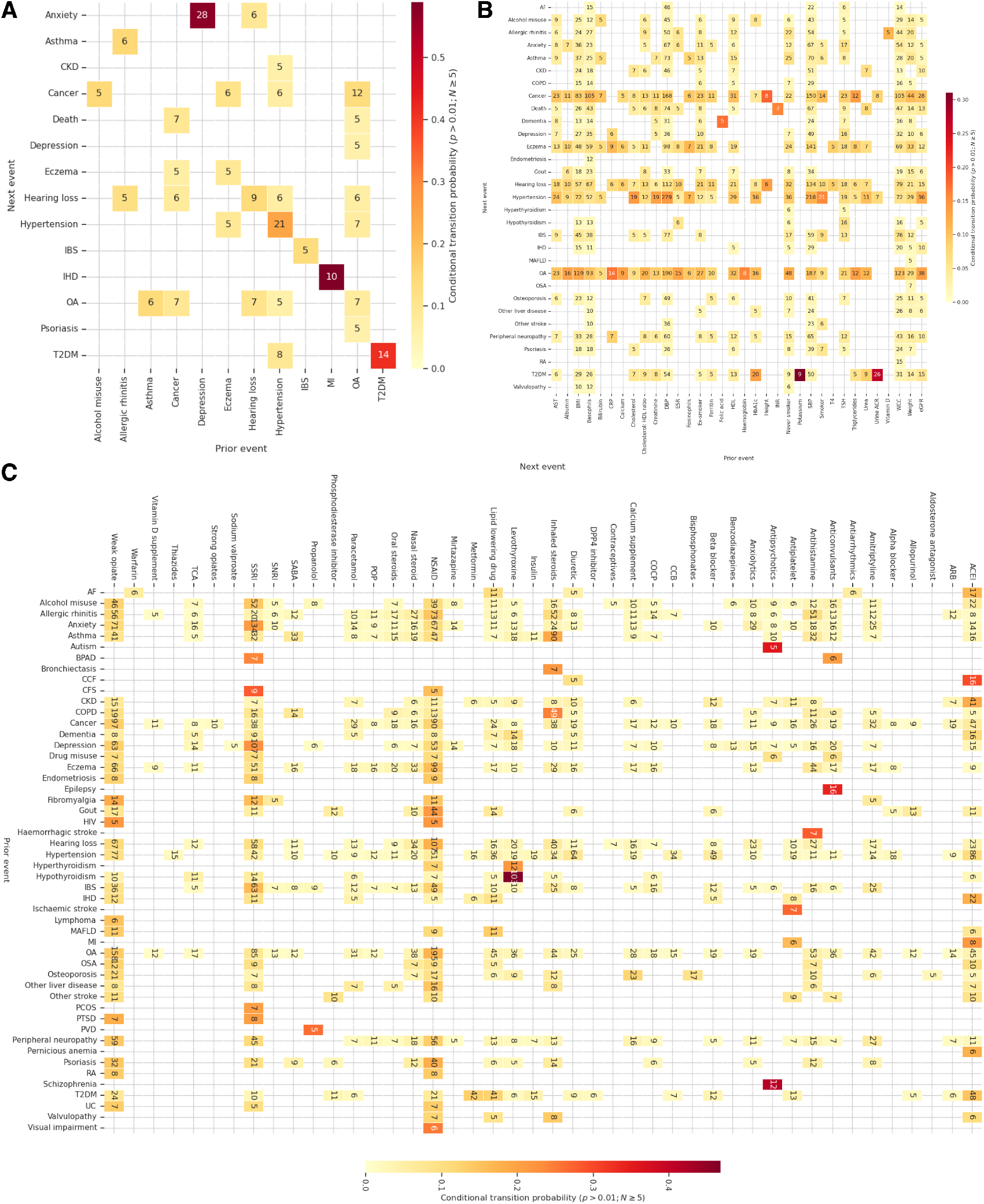
Forecasting known clinical associations. Given unseen patient histories from the pre-training cohort, we predict the next 3 events a patient may experience. From these we plot the transitional probability matrices for **A** a new diagnosis following a previous diagnosis, **B** a diagnosis follows an investigation and **C** medication follows a diagnosis. Colour scales indicate the SurvivEHR-derived conditional transitional probabilities while numbers indicate the observed occurrences during forecast generation. Transitions with fewer than 15 occurrences and probability below 0.05 are removed for clarity.

When we examined what SurvivEHR would predict as possible diagnoses following investigations (Figure 4B, Supplementary Figure S7), we observed that SurvivEHR predicts hypertension as a next event often after systolic and diastolic blood pressure (SBP/DBP) measurements as expected. However, preceding cholesterol, creatinine, eGFR measurements were also associated with hypertension diagnoses and these are routinely measured when there is clinical suspicion of hypertension. Weight, body mass index (BMI) and smoking status were also associated preceding events. We also found the expected transition that Type 2 Diabetes Mellitus (T2DM) diagnoses follow blood glucose (HbA1c) measurements was also recovered which is consistent with clinical practice in the UK [36].

Finally we examined which medications were forecast by SurvivEHR following a diagnosis (Figure 4C, Supplementary Figure S6). In the case of T2DM, these were metformin (a standard first-line treatment) and lipid-lowering and ACE inhibitors which are routinely prescribed to address the increased risk of cardiovascular and kidney disease in T2DM patients [36]. Schizophrenia was followed by antipsychotics. Epilepsy by anticonvulsants. Osteoporosis was followed by prescriptions of bisphosphonates - a gold standard, first line treatment - and calcium supplements which are supportive medications for the condition [37]. However, as an indication that SurvivEHR predictions are not *causal*, we see that osteoporosis was also associated with subsequent non-steroidal anti-inflammatory drugs (NSAID). NSAIDs are not indicated for osteoporosis itself but could be prescribed to manage associated (pain) symptoms or comorbid conditions. Similarly, visual impairment itself is not an indication for prescribing SSRIs (selective serotonin reuptake inhibitors). However, SurvivEHR’s forecasts have likely learnt that SSRIs may be prescribed in people with visual impairment if they have associated mental health conditions, such as depression or anxiety, which are common in individuals coping with vision loss.

Similarly, we also considered how risk profiles for individual patients change by varying different aspects of their history. To demonstrate this we curate a hypothetical female patient, born in 1963 and with Asian ethnicity. We model their history upon a real patient from the pre-training held-out cohort who did not experience any morbidities. Given this history of blood pressure, weight, blood test, smoking status monitoring, and previous anti-inflammatory drug medications, we can then modify this to transcribe our patient into three potential risk categories by varying their risk factors. In each case we mask any recorded weight values to ensure the record remains self-consistent. We define three categories: (i) low risk: a non-smoker, 120/80 mmHg blood pressure, and BMI of 24, (ii) moderate risk: an ex-smoker, 140/90 mmHg blood pressure, and BMI of 28 and (iii) high risk: a current smoker, 150/100 mmHg blood pressure, and BMI of 32. Given these histories, we can then forecast what their future risk for different events would be. Figure 5 shows that high-risk individuals with obesity are more likely to have body mass index measurements taken again than the other risk groups. While the risk of mortality, T2DM, heart failure and hypertension is also significantly increased as would be expected.

**Figure 5:**
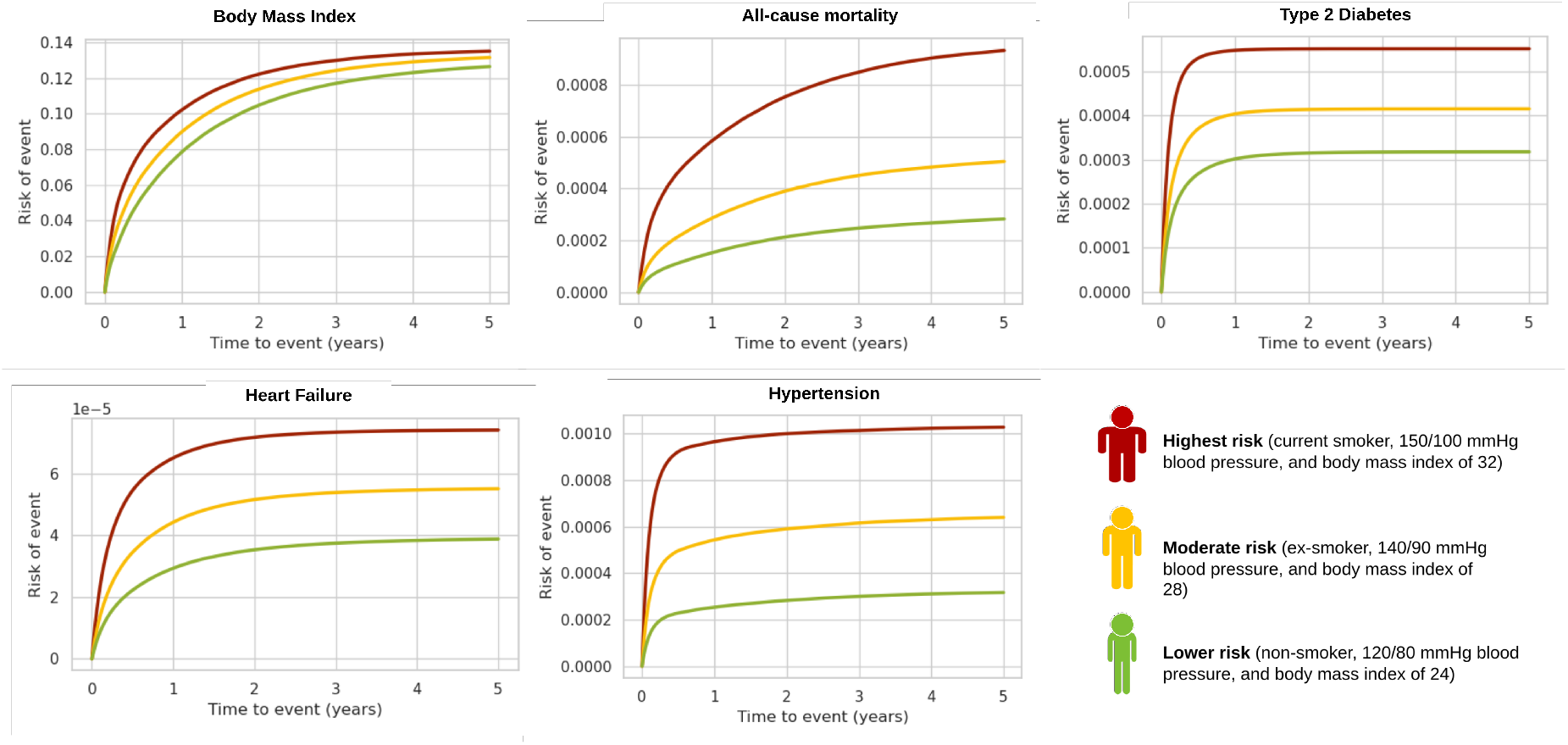
Illustrating SurvivEHR Predictive Behaviour. Next event risk prediction output for a variety of events (BMI measurement, all-cause mortality, type 2 diabetes mellitus, heart failure, hypertension) for three hypothetical patients with low, medium and high-risk health profiles. SurvivEHR output demonstrates the expected behaviour with higher risk factors.

These findings indicate that SurvivEHR is learning genuine clinical associations and risk factor relationships. Note that this experiment should not be considered causal prediction [38] since SurvivEHR only learns associative and not causal relationships.

### Clinical risk prediction and fine-tuning performance

We next evaluated SurvivEHR on two clinical risk prediction tasks, both aimed at assessing five-year risk following a diagnosis of type 2 diabetes (Figure 6A). The first task involved predicting the onset of hypertension – a classic single-outcome scenario. The second was a competing risk prediction of cardiovascular disease (CVD), which is inherently more complex due to the presence of multiple, mutually exclusive outcomes that collectively define CVD (see Methods).

**Figure 6:**
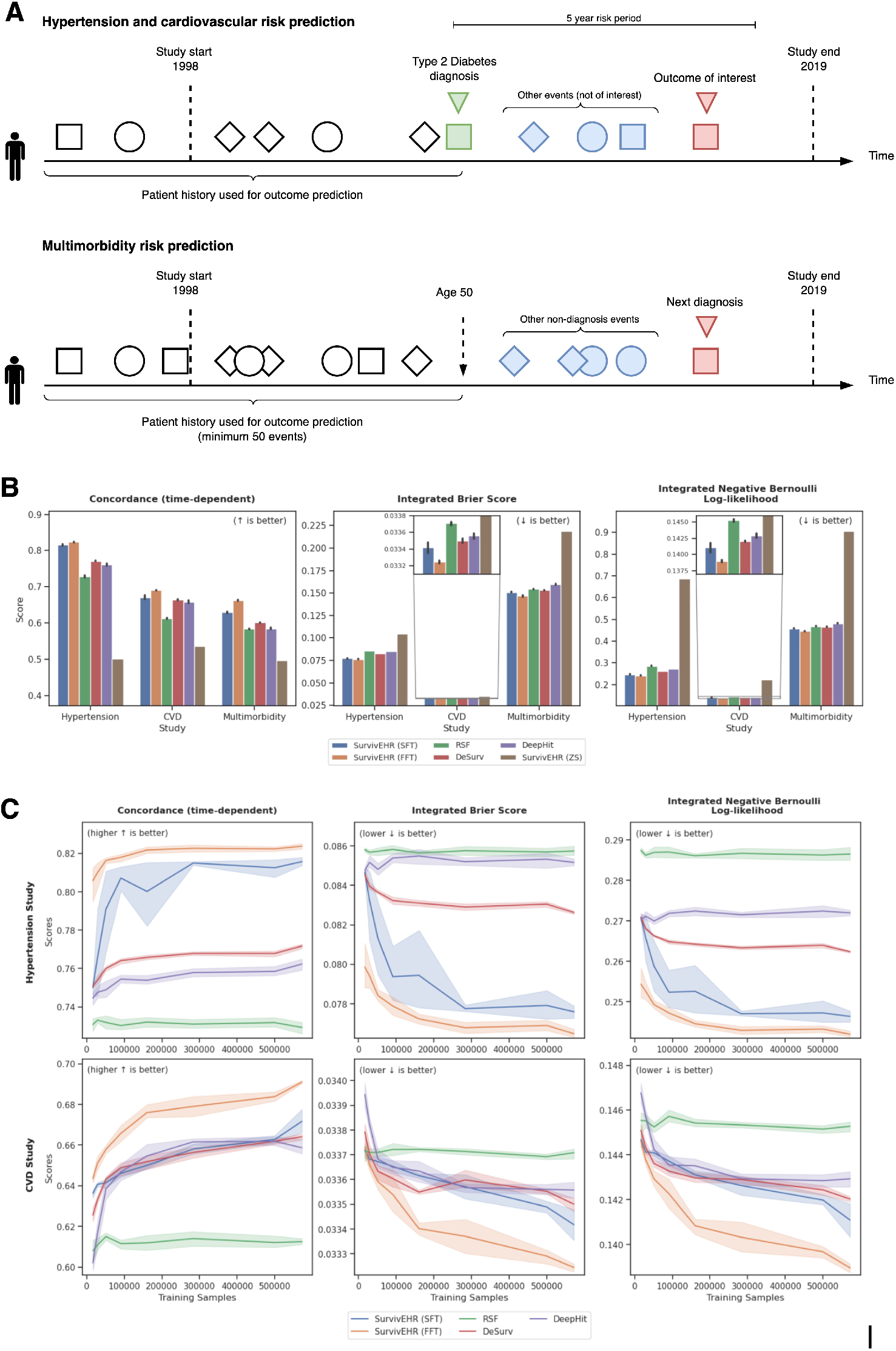
Fine-tuning prediction performance evaluation. **A** Fine-tuned models for retrospective cohort studies are trained and evaluated on contexts up to and including index date. Subsequent events are excluded until either (i) an outcome is observed or (ii) the last observation within the study period, which is then treated as a censored target. **B** Survival performance metrics for each cohort. **C** Ablation study of increasing cohort study population size.

For benchmarking predictive performance, we constructed three competing risk, time-to-event prediction models using ensemble and deep learning approaches (Random Survival Forests (RSF) [39], DeepHit and DeSurv [13]) using a cohort of 572,096 patients. We note that RSF, DeSurv and DeepHit use cross-sectional (most recent) inputs and not full patient histories. The alternative models (Table 1) that use longitudinal input do not provide competing risk capability.

We compared these models to predictions from two fine-tuned versions of SurvivEHR. The first was a scratch fine-tuned (SFT) version which uses the SurvivEHR architecture but was not pre-trained. The second version used full fine-tuning (FFT) which was first pre-trained before being fine-tuned for the five-year prediction task. This allows us to examine whether any performance improved is due simply to having a complex architecture (SFT) or if the pre-training adds information (FFT). We also tested the zero-shot (ZS) performance of SurvivEHR on this task by evaluating how well the pre-trained model can perform this task without any task-specific training.

We report three standard metrics, the time-dependent concordance measures how well a model is able to distinguish risk between patients who experience an event at different times; the Integrated Brier Score quantifies how well the model can accurately predict the probability of an event occurring over a period; whilst the Integrated Negative Binomial Log-Likelihood measures how well the model prediction matches the actual occurrence of the event.

Figure 6B (Table 2) shows that SurvivEHR-FFT achieves superior predictive performance to the other benchmarks on both tasks across all metrics for both prediction tasks. This demonstrates that pre-training provided additional prior information and that improved predictive performance did not arise purely due to increased model complexity and use of longitudinal input over RSF, DeSurv, DeepHit and SurvivEHR-SFT.

**Table 2.**
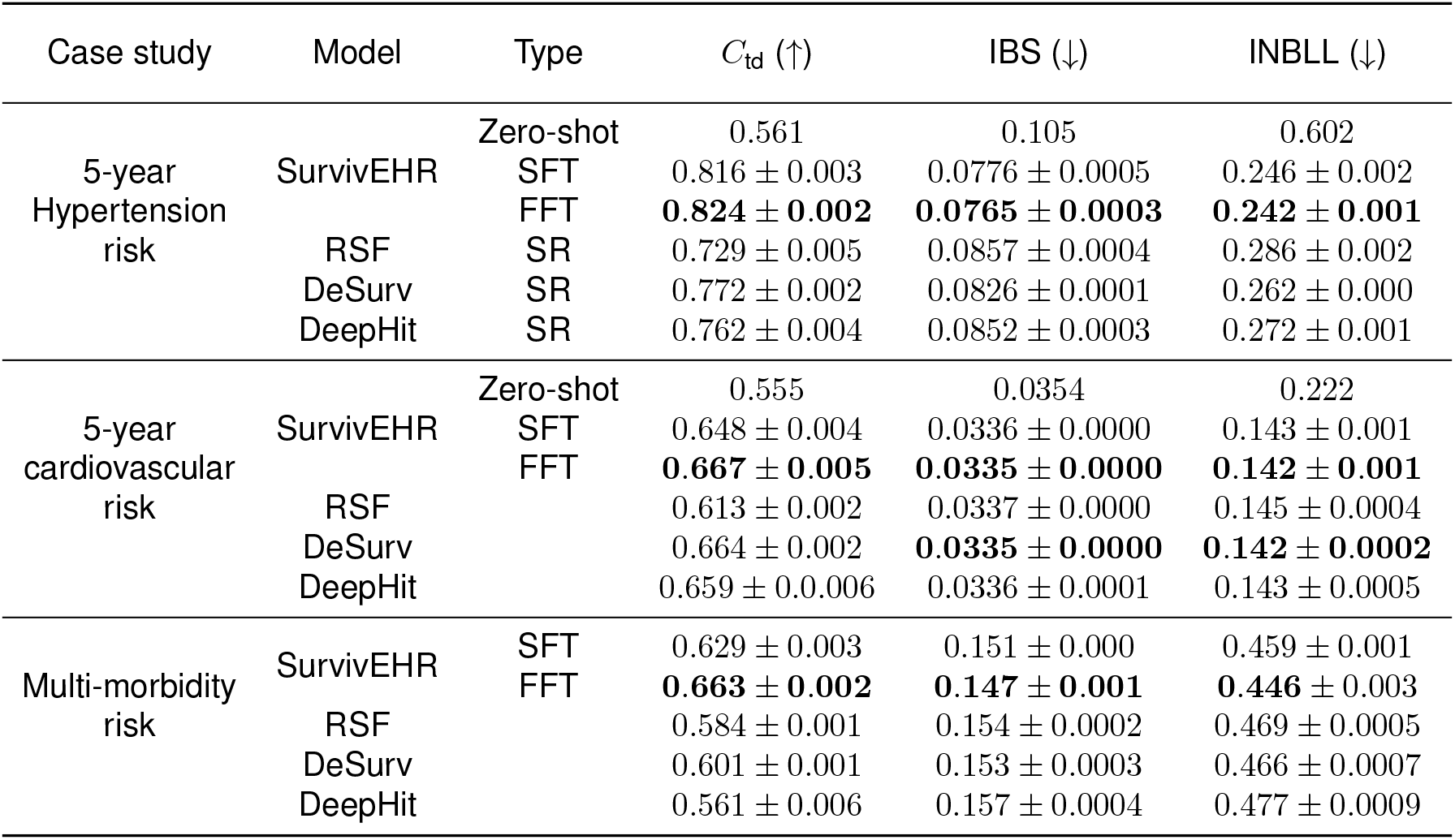
Clinical risk prediction model performance. Zero-shot indicates the evaluation of a pre-trained model, without any cohort specific training. Scratch Fine Tuning (SFT) indicates the CPM model was trained from scratch. Full Fine Tuning (FFT) indicates that we initialised training from the pre-trained model. Whilst single risk (SR), or competing risk (CR) indicates we replaced the pre-training survival head with a new survival model. The survival metrics (*C*_td_, IBS and INBLL) evaluate the model’s ability to predict the final outcome, and we report the average and 95% confidence interval over 5 random seeds.

These improvements were more evident if the patient cohort size is reduced from 572,096 patients to something substantially smaller. Figure 6C shows that when the cohort size is reduced, fine-tuning from a pre-trained model helps to maintain predictive performance and there are substantial benefits if the fine-tuning cohort size is less than 100,000 over the alternative models considered. Further, using a pre-trained model significantly improves model stability, with models trained from scratch leading to higher performance variability.

We next considered a more challenging risk prediction task of predicting the development of future co-morbidities at age 50 for patients with a heterogeneous multimorbid health background. Answering such questions is essential for proactive healthcare and effective disease management since co-morbid conditions significantly impact patient quality of life, healthcare costs, and mortality risk, making early identification a critical priority for public health. As before, SurvivEHR achieved superior predictive performance across all metrics for this more complex prediction task compared to the other baselines (Figure 6B, Table 2).

These results demonstrate that fine-tuning from a single pre-trained foundation model is able to enhance performance for different and variable predictive tasks. However, we note that zero-shot predictive performance is not attained by SurvivEHR and this can be attributed to the difference between the pre-training (next event) and evaluation (next outcome) objectives.

## Discussion

We have described a foundation model (SurvivEHR) that supports the development of clinical risk prediction models for patients with multiple long-term conditions. The foundation model leverages primary care EHRs that offer rich longitudinal trajectories that span decades of patient care, encompassing the full spectrum of health states from wellness monitoring through chronic disease management to end-of-life care. We exploit EHR data for a substantive proportion of the UK population to create a decoder-only transformer architecture that is pre=trained using a competing risk, time to next event objective.

Our experiments indicate that SurvivEHR can recapitulate known clinical associations and risk factor relationships, validating the fundamental premise that primary care EHRs contain learnable patient trajectories. Critically, these associations emerged without explicit programming or clinical rules, suggesting that the model architecture successfully learned the underlying epidemiological structure embedded within primary care practice patterns. These insights, captured in the embedding properties of SurvivEHR, can be exploited using fine-tuning to create more powerful clinical risk prediction models that utilise the insights gleaned from the extensive population records seen and captured by SurvivEHR.

The latter is an important benefit. Due to sensitivity and privacy issues, access to individual-level patient medical records. Access to large-scale real-world patient populations, like those provided by CPRD, are typically highly restricted and granted under limited conditions of use after extensive review and approval of an intended use plan. Pre-trained foundation models like SurvivEHR can provide an alternative approach for data sharing and model development by providing embedding functions pre-captured from individual-level data without the need to access or share the individual records themselves. As such, while our specific instance of SurvivEHR is trained only on a UK population and its most prevalent MLTCs, we envisage that similar models could be trained on other data sets particularly in different health systems which would account for variations in practice and clinical care guidelines. However, one issue of note is that we cannot provide full guarantees that the generative capability of SurvivEHR could not fully recapitulate real individual records and this currently places a limitation on model sharing. This is a general issue to be considered for all generative EHR models.

A limitation is that we have identified that SurvivEHR is not capable of zero-shot predictive performance. Fine-tuning is required to obtain competitive performance for specific risk prediction tasks. This limitation maybe inherent given the next event pre-training objective which limits the predictive horizons of the pre-trained model. We observed that this limited reliable forward generating capacity of the model to only a few steps. Future work could examine whether higher capacity model architectures can extend beyond next event prediction. However, this approach might not be effective in a primary care setting due to the diversity of clinical trajectories and the relatively sparsely recorded health indicators which limits predictive power. In addition, like other DL EHR prediction models, SurvivEHR is trained using associative learning only and cannot produce causal predictions. It is also subject to biases contained within the training data. Disentangling the many sources of bias and confounding that are contained within population-scale EHR data remains an open challenge.

We further note that SurvivEHR was constructed only from coded medical data and did not use potentially free-text data that exists in primary care records. Free text data is omitted from the research data provided by CPRD and the limitations and potential biases of this omission have been previously studied [40]. Future model developments could integrate free text and other data modalities though it should be noted that there is often limited availability of integrated population health data for research and a lack of computational and data infrastructure within clinical settings for the implementation of predictive analytics that might make use of such data.

In summary, SurvivEHR adds a distinctive contribution to a growing body of work in foundation models for EHR by providing one of the first decoder-only foundation models to learn full time-to-event distributions directly from a UK primary care EHR database (CPRD). We focused specifically on modelling 74 long-term conditions (and associated tests and medication classes) in a primary care setting in order to address a major challenge for global health systems that is currently under-represented in artificial intelligence research applied to electronic health records analysis.

## Methods

### Clinical Practice Research Datalink

We used the Clinical Practice Research Datalink, which contains longitudinal primary care data from a network of over 2,000 general practices in the UK that use EMIS Web^®^ patient record software. Around 1 in 10 GP practices, with 10 million currently registered patients and another 30 million historical patient records contribute data to CPRD. CPRD is broadly representative of the population by age, sex, and ethnicity. It has been extensively validated and is considered as the most comprehensive longitudinal primary care database, with several large-scale epidemiological reports adding to its credibility. Access to CPRD data was approved as part of the “OPTIMising therapies, disease trajectories, and AI assisted clinical management for patients Living with complex multimorbidity” (OPTIMAL) study [31].

### Data pre-processing

CPRD data was extracted using DExtER [29]. Clinical observations, diagnoses and treatments are recorded as Read Version 2, SNOMED-CT, and EMIS Web^®^ clinical codes. Prevalent cases for all 74 long-term conditions were identified using disease-specific clinical codelists. Code lists for 81 classes of medications associated with management of the 74 conditions were identified from prescription data recorded as Prodcodes from the Dictionary of Medicines and Devices (DM+D) codes, which are a subset of the SNOMED-CT terminology. We used 108 types of measurement and test (investigation) results, with values, reflecting routine blood tests such as haemoglobin, total cholesterol or serum creatinine levels, and clinical measurements such as blood pressure readings that are also routinely recorded in CPRD (Supplementary Table S1). Code lists can be obtained from Github (https://github.com/aditya02acharya/optimal_data).

The list of conditions, investigations and medications to be included was determined through an iterative process of discussion amongst a group of clinical experts including GPs, public health consultants and geriatricians, with further guidance from our patient advisory group. The starting point for these discussions was the findings of a seminal Delphi study that identified 59 key conditions that should be included in research for patients with MLTCs [30]. Our systematic process of developing clinical codelists and drug codelists using the DExtER code builder tool has been described in detail elsewhere [31]. We considered patients from England with at least 12 months of acceptable data recording prior to the index date (1 January 2005). Acceptable data was determined using the “acceptable patient flag” data quality measure provided by CPRD: (consistent recording of events including date of birth, practice registration date and transfer out date, and valid age and sex). There were 26.5 million patients across 1,478 practices meeting our inclusion criteria.

Pre-processing was performed using FastEHR, a custom Python toolkit that implements a high-throughput extract-transform-load machine learning data pipeline for EHR data extracts. It ingests raw data, builds an indexed SQLite store, and then uses the Polars library to stream large tables through memory-efficient transformations and filters. The pipeline handles: tokenisation; data cleaning, such as outlier removal and de-duplication; and allows custom dataset and dataloader curation for pre-training and fine-tuning. Users can specify flexible inclusion, exclusion, indexing and pre-processing criteria suitable for both self-supervised pre-training and supervised fine-tuning, whilst controlling for general practice- and dataset-level data leakage. In our workflow, FastEHR was developed as the data pre-processing backbone preceding model training with the SurvivEHR foundation model and is made available alongside the SurvivEHR package.

Following pre-processing with FastEHR, we were left with 51,003,640 diagnoses, 3,970,984,804 medications, and 3,533,426,831 investigations, of which 3,364,437,065 included the accompanying investigation result (Supplementary Table S2). Of these records, we employed a 90-5-5 site-level training, validation and test random split, dividing patients by general practices in England to avoid data leakage across practice. This left 23,613,894 patients from 1,330 practices in the training cohort, 1,426,714 patients from 74 practices in the validation cohort, and 1,508,320 patients from 74 practices in the test cohort. Statistics of the data extracted through DExtER and bounds used for internal scaling and outlier removal by FastEHR can be found in the Supplementary Information (Supplementary Tables S3–6).

### Principles of Model Design

Deep learning-based prediction models vary by architecture but broadly fall under three categories: encoder-only, encoder-decoder and decoder-only. Encoder only models (e.g., Med-BERT [41], BEHRT [42]) use masked language modeling or contrastive learning to learn contextual representations and are well-suited for classification and embedding tasks. Encoder-decoder models (e.g. TransformEHR [43]) support sequence-to-sequence tasks, are similarly trained, and are effective for summarization, question answering, and multi-task learning via prompting. Whilst decoder only models (e.g. MedGPT [44]) use autoregressive training for generation, and scale well for large modelling and generative applications. A summary of existing deep learning based clinical risk prediction models is given in Table 1. For brevity, we include only those clinical foundation models built for structured clinical codes rather than other modalities including free text and images.

In this work, we are interested in decoder-only approaches for causal next-event prediction. Many causal next-event prediction architectures have been used as base structures for EHR models. Each of these approaches typically adopt a two-stage training approach. Pretraining is first performed on large-scale longitudinal EHRs using self-supervised objectives to learn general-purpose representations of clinical features. This is then followed by fine-tuning on downstream clinical tasks such as diagnosis prediction, treatment recommendation, or risk stratification. However, by using architectures originally designed for language modelling, many EHR-based prediction models are limited to classifying the next most likely diagnosis or outcome at the next patient visit, and are either unable to provide a prediction for when this event will occur, or are only able to provide a point prediction.

Consequently, without explicit modelling of event timing, these models cannot properly account for censoring and important temporal risk information is lost, which can limit the clinical utility. For example, they cannot be used to directly answer questions such as “What is this patient’s risk of heart failure in the next X months?” for any arbitrary future time period. While previous models often include time or age-based embeddings to model the temporal order and distancing between events, this is not the same as learning time-to-event distribution models which can automatically account for irregular and variable follow-up durations. Recently, this limitation has been overcome with MOTOR [45], a self-supervised, time-to-event (TTE) foundation model but this is limited to approximations of time-to-event distributions using Piecewise Exponential Artificial Neural Networks.

The objective of SurvivEHR was to predict the time to the next recorded primary care event given a stream of previous primary care encounters and events (the patient history). This problem is highly related to generative (causal) modelling and is a form of self-supervised learning since histories can be retrospectively partitioned to set up supervised prediction tasks. Generative models built on this underlying task have become extremely popular in recent years due to their strong zero-shot generalisation [46], apparent emergent capabilities [47, 48] and scaling laws [49]. GPT models learn representations of sequences and, regardless of the form of these sequences, require each element to be vectorised. This is typically achieved through a process known as tokenisation and is then usually followed by a projection of each token. An attention mechanism then captures dependencies between tokens, allowing the model to weigh the importance of each token relative to others, regardless of their position in the sequence. This process allows GPT models to capture contextual relationships and generate coherent outputs by predicting the next token based on prior ones, effectively learning patterns and relationships within the sequence over time.

For each patient, we have a temporal sequence of primary care events that occur at irregular intervals. Each element of this sequence is broken down into a tuple of three items: a categorical event index, corresponding to a unique combination of medical codes; any attached numerical value, such as an investigation result; and finally the event time, in days to/since birth. The causal modelling task is to predict the next event and the associated time-to-event distribution, given the baseline covariates and the sequence of previous records. For instance, this may be predicting the risk of the next event being a diagnosis of substance abuse, and the corresponding time-to-event. In this example, no corresponding target value exists. This is visually depicted in Figure 1B.

### Model architecture implementation

#### Event embeddings

Here we outline our process for vectorising each electronic health record event. Each event contains an event category (comprising a collection of medical codes), an optional associated value, and continuous time since birth. In doing this, our task is to learn a numerical representation for each record. This is split into three independent embedding maps for: the static baseline covariates; a split embedding encoding the categorical event and any associated value; and the positional information of time of recording. We project each electronic health record event onto a 384-dimensional latent space.

##### Static embedding

For each event in a patient’s lifetime, we include a representation of their baseline covariates. These include their sex, ethnicity, year of birth, and postcode derived index of multiple deprivation^3^, and are considered fixed for the entirety of the patient’s lifetime. To allow the inclusion of missing records, we use one-hot encoding for each variable (with the exception of year of birth), despite the ordinal nature of the index of multiple deprivation.

Although these variables do not change over time, we additively include their embedding with each event to provide context consistently across the entire sequence. This is achieved by applying a linear projection to the concatenated encoded variables

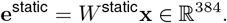

##### Split valued-event embedding

Each electronic health record event, which occur over the course of a patient’s lifetime, are included through a *split* embedding. Here, we obtain a numerical representation of each recorded pair of event category *p_c_*, and optional event value *v_c_*, at the *c*-th event of a patient’s timeline. Towards this end, we employ a split approach, composed of two embedding networks

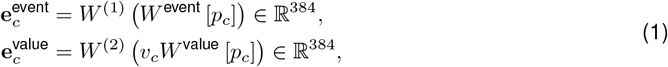

where *W* ^(·)^ [*p_c_*] are the rows of the embedding matrices *W* ^(·)^ corresponding to the token index *p_c_*. The associated values are encoded with a second (value-weighted) projection. Additionally, we observe that deviations of values from the population average contribute to the event embedding.

##### Positional embedding

By default the Transformer architecture is agnostic to the order at which events occur. As a consequence, it is typical to encode the positional information in each record embedding, encapsulated as the days to/since birth *t_c_* when the *c*-th event occurred.

There are many strategies for this, such as positional encodings, or positional embeddings [50, 51], which both which account for irregular time events. Within our experiments we use the original positional encoding of [50]. However, experiments with learnt positional embeddings showed no benefit for the increased computational cost:

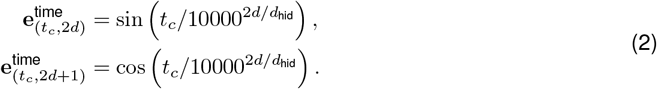

##### Combined event embedding

Each of these embeddings are additively combined to obtain the record embedding

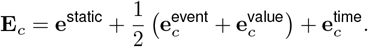

Putting these event embeddings up to event **E***_c_* into the Transformer provides us with a representation of a patients medical history denoted by **T***_c_* (Supplementary Figure S1).

#### Survival and value pre-training prediction

Causal Language Modelling predicts the next word in a sequence based solely on the preceding context encoded as **T***_i_*, following the causal structure of natural language generation. Here, our goal is to replace the typical black-box network which predicts next token logits with a new approach which instead predicts the full time-to-event survival distribution. This can then be used within the generative causal language modelling framework to infer a patient’s future sequence of health records.

Given a sequence of health records **r***_c_* = (*p_c_, v_c_*), occurring at irregular times *t_c_*, this translates to predicting the next event **r**_*c*+1_, and the time-to-event Δ*t*_*c*+1_ = *t*_*c*+1_ − *t*_*c*_ by providing accurate personalised cumulative distribution functions of the survival time. That is, given the history of a test subject, be able to provide an accurate estimate of the distribution over the survival time for each next event that may occur, and any value associated with this event.

##### Causal survival pre-training

Given a medical history encoded through **T***_c_*, survival analysis aims to predict the varying risk of a future event *p*_*c*+1_ (distinguishing event type), over the time-to-event Δ*t*_*c*+1_. A single risk approach assumes each of the possible next events can be considered in isolation, whilst competing risk assumes only one of the possible next events can occur in lieu of the others. Within the accompanying paper, we present results based upon competing risk pre-training.

##### Single-Risk with SurvivEHR

In the single risk setting we consider an outcome *k*_*c*+1_ ∈ [∅, 1}, where *k*_*c*+1_= 1 denotes that the following event, *p*_*c*+1_, belonged to some outcome set, and *k*_*c*+1_ = ∅ denotes right-censorship. For pre-training, this is a singleton set and we define 263 separate single risk models for each event in our vocabulary. For each model *m* = 1*, …,* 263 we have

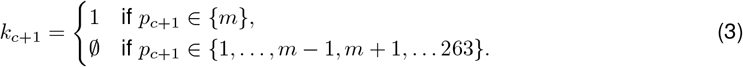

This is synonymous to 263 separate 1-vs-all single risk models. The goal is to learn a collection of models which are capable of providing accurate personalised prognostic cumulative distribution functions of the survival time for each event. That is, given a patients baseline and medical history, be able to provide an accurate estimate of the cumulative incidence function (CIF)

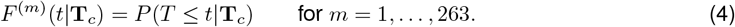

This encodes the probability of experiencing event *m* at a time *T*, by time t.

##### Competing-Risk with SurvivEHR

Alternatively, we consider the case of competing risks. In the causal setting, we instead have *k*_*c*+1_ ∈ [1*, …,* 263}. By default, the self-supervised learning is well suited for the competing risk case, as each subsequent event may be only one of the considered events (under the assumption that event times follow a Poisson process).

Further, this means no null event ∅ is typically needed during pre-training.^4^ In the case of pre-training for competing risks, the CIF associated with each event type is defined as

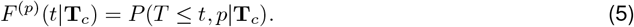

This is related to the CIF for the overall survival time (until any next event) by

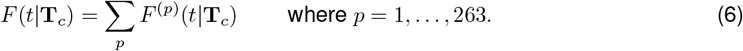

In each case, the Transformer’s hidden states, **T***_c_*, are used as input to a deep survival model to predict survival outcomes. For this we choose to use DeSurv [13], a flexible neural network based approach for modeling survival distributions in continuous time. However, alternatives such as Cox-based survival models (e.g. PyCox [52]) or alternative neural approaches (e.g. SumoNet [53], Neural Fine-Gray [54]) could also be used.

##### Generation

In both single and competing risk cases, our survival heads predict a cumulative density for each separate event over a fixed time grid. Sampling the time-to-event within each is achieved through a two-stage approach. First, we sample which event will occur next with a probability proportional to the area under the cumulative density over the 5-year period. Secondly, we sample the time to event using inverse transform sampling.

#### Causal value prediction

For each valued-event (such as those events *p* for which a value *v* may exist) we define a probabilistic regression layer predicting a Gaussian mean and standard deviation, conditional upon the hidden representation **T***_i_*. The resultant log-likelihood is then combined to the loss additively. This approach comes with a limitation that predicted values share the same distribution across all future times. Consequently, we will not observe a pharmacological onset time. This simplification is similarly made in existing state-of-the-art approaches, and is left as future work.

#### Hyperparameters

Model hyperparameters used for training are given in Supplementary Table S7.

### Pre-training evaluation metrics

#### Survival metrics

Survival metrics evaluate the performance of models predicting time-to-event outcomes, whilst accounting for unique challenges like censoring. Key metrics include the concordance index (c-index), which assesses the model’s ability to correctly rank relative risks between patients, the Integrated Brier Score (IBS), measuring time-dependent prediction accuracy, and the Negative Bernoulli Log-Likelihood (NBLL), which quantifies the likelihood of observed survival times given model predictions. These metrics enable rigorous comparisons and validation of survival models, ensuring robust and reliable predictions. In part, they are able to achieve this by accounting for censored samples, which represent incomplete but valuable information about survival times. Whilst improper handling of censored samples leads to a poor reflection of the true predictive performance of survival models in real-world scenarios where complete data is rare.

Concordance metrics, for example, quantify the consistency between the relative predicted risk between patients, compared to the observed risk. If patient *i* and *j* both experienced an event, the concordance is higher if the model correctly predicts which patient experiences the event first. This *intra*-event metric is individually calculated for each event across concordant pairs of patients. Many estimates of this metric for survival analysis exist, with the simplest being the Harrell’s c-index which counts the proportion of concordant pairs

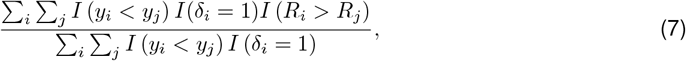

where *i* and *j* index patient pairs, *y_i_* is the minimum of the survival time *t_i_* and censoring time *u_i_, δ* = *I* (*t_i_ ≤ u_i_*), and *R_i_* is an estimated risk score under some model [55].

#### Next event prediction performance

However, event sparsity in electronic health records makes within-event pair construction computationally impractical. For example, rare events such as Addison’s disease occurred at a frequency of 6,691 diagnoses in 7.6 billion events, making within batch computation practically infeasible. Instead, we propose a new metric based upon the discrimination approach of Harrell’s c-index, but instead considering between event discrimination. This leads to a metric similar to those found in recommender systems, such as the Mean Reciprocal Rank.

##### Inter-event concordance (IEC)

We employ a new metric to assess a pre-trained model’s capacity to order survival risk between events in the self-supervised setting. We are interested in developing an *inter* -event metric that is capable of determining consistency in the relative predicted risk of all possible next events, compared to the observed true next event.

In this settings, rather than quantify the ability to capture relative risks between pairs of patients, we quantify the accuracy of the pre-trained model in correctly ranking the risks of the next true event, across all possible next events. In doing this, we obtain a discrimination score for each individual event category quantifying how accurately the model is able to rank the relative risk.

We now update the above index notation: given all cases where an event *i* ∈ [1*, …,* 263} occurred next, we derive a score quantifying how well the risk of this event was ranked across all 263 possible predicted next events, *j*

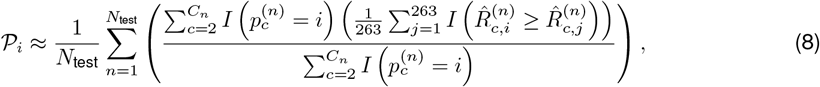

where 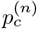 is the index of the event experienced by patient *n* after experiencing *c* prior events. Similarly, 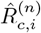 is the predicted estimated risk score for the next event *i*, for patient *n* after observing the *c* prior events. For the risk score, we use the restricted mean survival time (RMST) calculated over a 5 year period. This is marginalised over both the number of patients *N*_test_, and each new event the patient experienced *C_n_*. In practice, we obtain an unbiased approximation of this by sub-sampling.

Through this approach we can quantify model alignment with the training objective, identify patterns that the model predicts confidently and highlight potential biases or weaknesses in its learned representations. This approach is particularly useful for understanding event-level prediction fidelity and for comparing the model’s performance across various pre-training datasets and tasks. Such metrics based on the Harrell’s c-index [56] are known to be a function of the censoring distribution [57, 58], inflating the scores of more prevalent events. This is similarly true for our proposed metric and consequently we compare our model to a baseline score which is obtained by following a prevalence only prognostic approach, in which the risk score 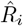 is equal to the total number of events *i* in the population.

The *IEC* scores for each medication and investigation are shown in **??**. These are calculated across every new event observed within each patients history for a subset of the unseen held-out patients. In each of these, some events may be missing due to sub-sampling.

#### Multi-step forecasting

We can use the Inter-event Concordance metric to evaluate how performance degrades as we look further ahead by updating Equation 8. Retaining the notation for event index *i* ∈ [1*, …,* 263}, we derive a score quantifying how well the risk of an event *t*-steps in the future is ranked across all 263 possible future events, *j*

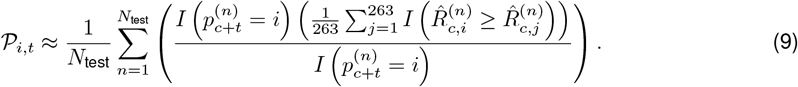

Here, *c* is chosen to be the index of the event *t* steps prior to each patient’s last observation, such that *c* + *t* is the number of records in the timeline of patient *n*. We do not calculate the score over the entire patient trajectory, but include the medical history up to *t*-steps before their final observation, and test the ability to predict the final observed event. Consequently, 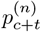 is the final event category experienced by patient *n*. We then evaluate how well the model was able to predict the risk of this event multiple steps in the future, despite being trained to predict 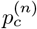.

### Fine-tuning experiments

In our fine-tuning experiments, we consider the downstream task of predicting the risk of a future outcome from a patient history up to an index date. Throughout all fine-tuning examples, we increase the context window to 512, prepend any previous diagnoses which may otherwise be lost from the context window, and remove repeated events. This ensured that all health conditions that could have lifelong implications are included for prediction. All benchmark examples are given full patient histories, and therefore are not limited by context length.

We considered two approaches to fine-tuning. The first we call supervised fine-tuning from scratch (SFT) in which the supervised model is trained from randomly initialized parameters. While in the second, we perform supervised full fine-tuning from the pre-trained model (FFT). This allowed us to distinguish between learning due to the model architecture and that which occurs through transfer learning. Additionally, we benchmark against a number of alternative methods to predict the 5-year risk using Survival Random Forests [39], DeepHit [12], and DeSurv [13]. For each we take a cross-section of the full patient history, including baseline covariates, all diagnoses, medications and investigations. This is composed of a concatenation of the baseline covariates and a binary vector, where cases where an event occurred before an index date are marked as 1, and 0 otherwise. Note that we could not include approaches (such as BEHRT) which process longitudinal input data but only provide single-risk, binary classification output for comparison.

We fine-tune on two different cohorts. For the first cohort we consider patients with type 2 diabetes mellitus and indexing at this diagnosis. After applying our inclusion criteria we are left with 572,096 patients, with an additional 35,758 reserved for validation and 33,280 for testing. Within this cohort we consider both single- and competing-risk tasks of predicting the 5-year risk of hypertension and cardiovascular disease (CVD) respectively. We took CVD to be the competing-risk of ischemic heart disease (including myocardial infarction) and stroke (including ischaemic stroke, haemorrhage, and from unspecified causes). For our second cohort we considered patients with pre-existing multimorbidity, indexing at age 50 and using a random sample of 20,000 patients for training.

A full outline of the inclusion criteria for each study is provided in the Supplementary Information.

## Code release policy, data and model availability

The full source code used to pre-process, train, evaluate, and reproduce our experiments is available at http://github.com/cwlgadd/FastEHR and http://github.com/cwlgadd/SurvivEHR under an opensource licence. As the proposed SurvivEHR model uses generative AI, we are not able to distribute the trained model weights to avoid compromising patient privacy and contravening data-sharing agreements as we cannot guarantee that exact copies of real records could not be reproduced by the model. We provide detailed instructions to allow others to retrain the model from scratch on appropriately licensed data. Data for this project was made available by CPRD under study reference ID 21_000683 (https://www.cprd.com/approved-studies/optimising-therapies-and-disease-trajectories-patients-living-complex). Raw data from the study are not publicly available. Data for the study were obtained under licence from CPRD; pseudonymised patient data are available from CPRD subject to Research Data Governance approval; see https://www.cprd.com/how-access-cprd-data for more information. Codelists for CPRD data extraction can be found at http://github.com/THINKINGGroup/phenotypes.

## Acknowledgements

This study/project is funded by the National Institute for Health Research (NIHR) Intelligence under its programme Artificial Intelligence for Multiple Long-Term Conditions under the title “OPTIMising therapies, disease trajectories, and AI assisted clinical management for patients Living with complex multimorbidity” (OPTIMAL study) under Award ID: NIHR202632 (https://fundingawards.nihr.ac.uk/award/NIHR202632). The views expressed are those of the author(s) and not necessarily those of the NIHR or the Department of Health and Social Care. Additional funding was provided by the UK Engineering and Physical Sciences Research Council (Ref: EP/Y018192/1). Christopher Yau is supported by an UKRI Turing AI Acceleration Fellowship (Ref: EP/V023233/1).

The OPTIMAL collaborative consists of the following members: Rebecca Birch, Marco Canducci, Dominic Danks, Alexander d’Elia, Alastair Denniston, Sarah Flanagan, Suzy Gallier, Naijie Guan, Xin Guan, Imane Guellil, Georgios Gkoutos, Shamil Haroon, Eleanor Hathaway, Louise Jackson, Janet Lord, Zeinab Majid, Tom Marshall, George Morris, Charlotte Owen, Elizabeth Sapey, Chris Sainsbury, Charlotte Spurway, Peter Tino, Steven Wambua (all University of Birmingham); Amaya Azcoaga-Lorenzo, Colin McCowan, Luciana Rocha Pedro, Muhammad Usman (all University of St Andrews); Natalia Hong, Sara Matijevic, Kaspar Martens (all University of Oxford); and Tim Williams, Puja Myles (MHRA).

We are grateful to the OPTIMAL patient advisory group, which included Bob Jasper, Emily Lam, Jenny Negus, Gillian Richards, Lynne Wright, Christine Michael and others. Their diverse perspectives and extensive lived experience of long-term conditions have been vital to this work. The patient advisory group were involved in reviewing the study design and development of the list of conditions for inclusion. They were also involved in interpretation of the study results, strengths, and limitations.

We are also thankful for the support of the OPTIMAL Research Steering Group: Professor John Gladman (Chair, University of Nottingham), Dr Philip Bell (Patient and Public Representative), Dr Derrick Bennett (University of Oxford), Professor Paramjit Gill (Professor of General Practice), Professor Claire Steves (Kings College London) and Professor Reecha Sofat (University of Liverpool).

## Author Contributions Statement

CG, KN and CY contributed to the conceptualisation of the work. CG designed experiments and summarised findings under the guidance of CY. JC and TJ provided clinical support for the research. CG provided support for the development of the methodology. AA, CG and KG prepared the data. FC and KN developed the research protocol. LF led patient engagement and involvement activities. CG prepared the original draft, and all authors participated in revising the article. TJ, KN and CY provided overall management and obtained funding for the project.

## Footnotes

1 We searched PubMed for relevant studies using the terms (“primary care” AND “deep learning” AND “electronic health record”) and found only 12 total results of which most were not relevant.

2 We only predict the next three events since previous analysis shows breakdown after four steps.

3 As patient records between different general practices cannot be linked within CPRD, each patient is tied to one registration and therefore any derived index of multiple deprivation scores remain constant throughout the patient's registration.

4 Though semantically we can consider vocabulary truncation (in which events of low frequency are excluded) to form this empty set.

