## Supplementary Information for "SurvivEHR: a competing risks, time-to-event foundation model for multiple long-term conditions from primary care electronic health records"

### A Data processing

#### A.1 DExtER

CPRD Aurum data is extracted according to a pre-specified study protocol using DExtER [1]. Under this protocol, DExtER extracts 74, 81 and 108 collections of diagnosis, medication and investigation codes, respectively. These are outlined in Table S1, where we additionally show the corresponding labels used for analysis and plotting. The total number of records extracted by DExtER is shown in Table S2.

##### A.1.1 Code list

The list of conditions, medications and investigation to be included was determined through an iterative process of discussion amongst a group of clinical experts including general practitioners, public health consultants and geriatricians, with further guidance from our patient advisory group. The starting point for these discussions was the findings of a seminal Delphi study that identified 59 key conditions that should be included in research for patients with multiple long term conditions (MLTCs). Our systematic process of developing clinical codelists and drug codelists using the DExtER codebuilder tool has been described in detail elsewhere. Codelists can be found at `github.com/THINKINGGroup/phenotypes`.

##### A.1.2 Summary statistics

Summary statistics for the data extracted using DExtER, before any pre-processing is performed, are given in Tables S3–S5. Count refers to the total number of examples extracted from CPRD. For investigations, for which we also have numerical observations, we also report the mean, minimum, and maximum statistics across observations. Additionally, we report the number of occurrences when an event occurred but the associated value was unavailable.

These statistics are obtained before any pre-processing steps are applied and demonstrate the irregular structure of the raw data, including both sparsity and wide variation in numerical recording. As a consequence of this we performed extensive pre-processing, including outlier removal outlined in subsubsection A.2.2, and chose to mask poorly recorded values relating to smoking status.

#### A.2 FastEHR

We developed FastEHR, an open-source Python package, to construct high-throughput machine learning datasets for structured electronic health records (`github.com/cwlgadd/FastEHR/`). Upon extraction of raw data by DExtER, we built an indexed, single-file database using SQLite which can be used to collate patient information and collect meta information used for pre-processing and reporting. This is then used to build flexible datasets that allow for different training objectives, study inclusions, cohort curation, and different outcomes.

FastEHR supports scalable preprocessing via Polars, patient- and practice-level inclusion criteria, configurable cohort splits, and different tokenization strategies. Additionally, FastEHR outputs model-ready datasets in Parquet format alongside metadata for reproducibility, and provides data loaders compatible with PyTorch, making it an efficient backbone for large-scale EHR model development.

##### A.2.1 Tokenisation

We employed whole-word tokenisation, where each token corresponds to an event category composed of the collection of medical codes for diagnoses, medications, or investigations extracted using DExtER. The full list of these event categories, their corresponding token index, and the short format names used for analysis and plotting are shown in Table S1.

The first two tokens are reserved for internal padding ("PAD") and an unknown ("UNK") token. Whilst unused in our experiments, the unknown token is designed to encapsulate any out-of-vocabulary event categories, flagging them so the model can still process sequences containing unknown events, even though it lacks a dedicated embedding for that item.

##### A.2.2 Outlier filtering and scaling

Feasible lower ($F_L$) and upper ($F_U$) outlier limits were chosen by clinical experts. For those values where clinician-derived limits were not available, we instead derived empirical bounds, defined by $F_L = Q_1 - 1.5 * IQR$ and $F_U = Q_3 + 1.5 * IQR$, where $Q_1$ and $Q_3$ are lower and upper quantiles, and IQR is the inter-quantile range. Quantiles were approximated by sub-sampling with t-Digest [2]. The limits used are given in Table S6.

Investigations whose values lay beyond these limits had their values removed during pre-processing and were subsequently treated as missing. In these cases, the token was still retained and consequently the model remains aware that the investigation occurred, but not what the investigation result was.

Internal scaling was performed on the remaining recorded values using min–max scaling on these limits. Motivated by our choice of embedding architecture, we chose to centre our standardisation at zero.

##### A.2.3 Missing data

Missingness is a common characteristic of electronic health records, particularly for investigation codes where numerical values may be absent or improperly recorded. We distinguish between missing values in the input (predictor) and target (self-supervised or supervised label) contexts.

For missing input investigation results we apply a simple imputation strategy. After applying the outlier removal and scaling procedures described above, missing values are set to zero, which corresponds to imputing the midpoint of the interquartile range, $\frac{Q_1+Q_3}{2}$.

For missing target values, we retain the remainder of the patient record but exclude the missing values from contributing to the loss function during training. This allows the model to learn from partially observed outcomes.

We are unable to account for cases where an event may have occurred but the medical code was not recorded in the patient's records. This is a consequence of their absence being unobservable and thus indistinguishable from true non-occurrence. However, we note that many of these cases will exist and adds to the challenging nature of primary care data.

### B Pre-training performance evaluation

#### B.1 Representation Learning and demographic bias

During pre-training SurvivEHR learns to produce representations that summarise patient histories. Here we explore this concept space. Using patients from the pre-training cohort, we explore each patient's timeline up to each diagnosis they experienced. For example, given a patient who has experienced diagnoses of anxiety, depression, and arthritis, we extract three trajectories up to and including each of these events.

First, we explore the effect of static covariates on the latent embedding. In Figure S3 we plot the UMAP latent embedding $\mathbf{T}_c$ up to and including each of their diagnosis events. In Figures S3a–S3d, we label by different static covariates: sex, ethnicity, deprivation level given by their Index of Multiple Deprivation, and year of birth. The only clear associations were sex and birth year, as both will be significant predictive risk factors in most health conditions. Additionally, in Figure S3e and S3f we plot the same embedding labelled by the number of historical morbidities, and the last seen diagnoses, respectively. For the former we see no clear pattern emerge. For the latter we see some emergent structure, including a clear cluster of eczema diagnoses that is neighboured by asthma diagnoses. However, this is explained by the birth year bias, where each can be explained by the age of the population in this area of the concept space. Additionally, we see a separate cluster of patients who died, but this is explained by this event marking the end of any patient records.

We continue by exploring the concept space of the same patients, but with the static covariate contributions $\mathbf{e}^{\text{static}}$ set to zero. These are shown in Figure S4. As before, we plot the concept space labelled by the number of morbidities and the last seen diagnosis. Here we again see no clear structure when labelled by the number of historical morbidities. However, when labelled by last seen diagnosis we see some further structure emerge. However, we note that there remains a lack of distinctive sub-structures.

#### B.2 SurvivEHR captures known clinical associations

Through the typical recursive strategy of predicting the next event, SurvivEHR is capable of forecasting future clinical trajectories. Taking a population of patients, we present SurvivEHR with each held-out patient's complete history and ask it to predict the next three clinical events. We then analyse the conditional transition matrix that captures how often events arise together. This is given by

$$P(\text{next} = i \mid \text{prior} = j) \;=\; \frac{N_{j\to i}}{\sum_k N_{j\to k}},$$

where $N_{j\to i}$ denotes the number of observed transitions from event $j$ to event $i$.

For the pre-training cohort, the resulting transition matrices are shown in Figures S5–S7 for observed transitions (A) from diagnoses to diagnoses; (B) from diagnosis to medication; and (C) from investigations to diagnosis. Cells based on less than $5$ observations or conditional probabilities less than $0.01$ are masked for clarity.

#### Tables

**Supplementary Table S1: SurvivEHR event vocabulary.** Each event category is composed of a collection of medical codes which are collated under the same token. Some categories, such as those recorded under different units, are combined for clarity during plotting.

| Event category | Plotting label | Token |
|---|---|---|
| PAD | - | 0 |
| UNK | - | 1 |
| ADDISONS_DISEASE | Addison’s | 2 |
| ADDISON_DISEASE | Addison’s | 6 |
| AF | AF | 81 |
| ALCOHOLMISUSE_V2 | Alcohol misuse | 94 |
| ALLCANCER_NOHAEM_NOBCC | Cancer | 104 |
| ALLERGICRHINITISCONJ | Allergic rhinitis | 123 |
| ALL_DEMENTIA | Dementia | 75 |
| ANXIETY | Anxiety | 126 |
| ANY_DEAFNESS_HEARING_LOSS_V2 | Hearing loss | 114 |
| AORTICANEURYSM_V2 | Aortic aneurysms | 30 |
| ASTHMA_PUSHASTHMA | Asthma | 134 |
| ATOPICECZEMA | Eczema | 136 |
| AUTISM | Autism | 39 |
| BIPOLAR | BPAD | 31 |
| BRONCHIECTASIS | Bronchiectasis | 32 |
| CHRONICFATIGUESYNDROMEMM_V2 | CFS | 26 |
| CHRONIC_LIVER_DISEASE_ALCOHOL | Alcoholic liver disease | 19 |
| CKDSTAGE3TO5 | CKD | 93 |
| COPD | COPD | 83 |
| CROHNS_DISEASE | CD | 23 |
| CYSTICFIBROSIS | CF | 3 |
| DEATH | Death | 106 |
| DEPRESSION | Depression | 133 |
| DOWNSSYNDROME | Down syndrome | 7 |
| EATINGDISORDERS | Eating disorders | 44 |
| ENDOMETRIOSIS_ADENOMYOSIS_V2 | Endometriosis | 48 |
| EPILEPSY | Epilepsy | 60 |
| FIBROMYALGIA | Fibromyalgia | 37 |
| GOUT | Gout | 79 |
| HAEMOCHROMATOSIS_V2 | Haemochromatosis | 8 |
| HF_V3 | CCF | 74 |
| HIVAIDS | HIV | 12 |
| HYPERTENSION | Hypertension | 129 |
| HYPERTHYROIDISM_V2 | Hyperthyroidism | 49 |
| HYPOTHYROIDISM_DRAFT_V1 | Hypothyroidism | 91 |
| IHDINCLUDINGMI_OPTIMALV2 | IHD | 95 |
| ILD_SH | ILD | 18 |
| ISCHAEMICSTROKE_V2 | Ischaemic stroke | 41 |
| LEUKAEMIA_PREVALENCEV2 | Leukaemia | 16 |
| LYMPHOMA_PREVALENCE_V2 | Lymphoma | 22 |
| MENIERESDISEASE | Ménière’s | 21 |
| MINFARCTION | MI | 67 |
| MS | MS | 14 |
| NAFLD_V2 | MAFLD | 40 |
| OSA | OSA | 52 |

*Continued on next page*

Supplementary Table S1 – continued from previous page

| Event category | Plotting label | Token |
|---|---|---|
| OSTEOARTHRITIS | OA | 120 |
| OSTEOPOROSIS | Osteoporosis | 78 |
| OTHER_CHRONIC_LIVER_DISEASE_OPTIMAL | Other liver disease | 57 |
| PAD_STRICT | PAD | 56 |
| PARKINSONS | PD | 29 |
| PERIPHERAL_NEUROPATHY | Peripheral neuropathy | 89 |
| PERNICIOUSANAEMIA | Pernicious anemia | 20 |
| PLASMACELL_NEOPLASM_V2 | Myeloma | 9 |
| PMRANDGCA | PMR | 45 |
| POLYCYSTIC_OVARIAN_SYNDROME_PCOS_V2 | PCOS | 58 |
| PREVALENT_IBS_V2 | IBS | 97 |
| PSORIASIS | Psoriasis | 82 |
| PSORIATICARTHRITIS2021 | Psoriatic arthritis | 13 |
| PTSDDIAGNOSIS | PTSD | 35 |
| PVD_V3 | PVD | 43 |
| RHEUMATOIDARTHRITIS | RA | 46 |
| SCHIZOPHRENIAMM_V2 | Schizophrenia | 34 |
| SICKLE_CELL_DISEASE_V2 | Sickle cell | 5 |
| SJOGRENSSYNDROME | Sjögren’s | 10 |
| STROKEUNSPECIFIED_V2 | Other stroke | 65 |
| STROKE_HAEMRGIC | Haemorrhagic stroke | 28 |
| SUBSTANCEMISUSE | Drug misuse | 70 |
| SYSTEMIC_LUPUS_ERYTHEMATOSUS | SLE | 11 |
| SYSTEMIC_SCLEROSIS | Systemic sclerosis | 4 |
| TYPE1DM | T1DM | 36 |
| TYPE2DIABETES | T2DM | 100 |
| ULCERATIVE_COLITIS | UC | 33 |
| VALVULARDISEASES_V2 | Valvulopathy | 62 |
| VISUAL_IMPAIRMENT | Visual impairment | 38 |
| ACE_Inhibitors_D2T | ACEI | 259 |
| AMD_DH_Donepezil | Donepezil | 142 |
| AMD_DH_Memantine | Memantine | 116 |
| ARBs_Luyuan | ARB | 221 |
| Acarbose_AURUM | Acarbose | 72 |
| AldosteroneAntagonist_D2T | Aldosterone antagonist | 155 |
| AllHIVdrugs_HIV | Antiretrovirals | 24 |
| All_AntiArrhythmics_D2T | Antiarrhythmics | 191 |
| All_Antiplatelets | Antiplatelet | 256 |
| All_Diuretics_D2T | Diuretic | 257 |
| All_Diuretics_ExclLactones_D2T | Diuretics (excl. lactones) | 258 |
| AlphaBlocker | Alpha blocker | 210 |
| Amantadine | Amantadine | 68 |
| Amitriptyline_optimal | Amitriptyline | 205 |
| Anticonvulsants_OPTIMAL | Anticonvulsants | 233 |
| Antihistamines_second_generation_H1_antagonists | Antihistamine | 207 |
| Antipsychotics_OPTIMAL | Antipsychotics | 196 |
| Anxiolytics_mumpredict | Anxiolytics | 198 |
| Aspirin_OPTIMAL | Aspirin | 253 |
| Benzodiazepines | Benzodiazepines | 213 |
| BetaBlockers_OPTIMAL | Beta blocker | 228 |
| Bisphosphonates_osteoporosis | Bisphosphonates | 195 |
| COCP_reg_contraception | COCP | 201 |

*Continued on next page*

Supplementary Table S1 – continued from previous page

| Event category | Plotting label | Token |
|---|---|---|
| COMT_Optimal | COMT inhibitor | 90 |
| CalciumChannelBlck_D2T | CCB | 250 |
| Calcium_supplements_Optimal | Calcium supplement | 216 |
| Carbamazepine_Optimal | Carbamazepine | 158 |
| DPP4inhibitors_OPTIMAL | DPP4 inhibitor | 156 |
| Dementia | Dementia | 151 |
| Digoxin_ | Digoxin | 174 |
| Direct_Acting_Oral_Anticoagulants_DOAC | DOAC | 164 |
| Fibrate_FenofibrateOnly | Fenofibrate | 115 |
| First_gen_H1_antihistamines | Antihistamine | 171 |
| GLP_1Aurum | GLP1 agonist | 121 |
| Gabapentin_Oral_OPTIMAL | Gabapentin | 175 |
| Galantamine_NEW | Galantamine | 88 |
| HydroxychloroquineAURUM_Zhaonan | Hydroxychloroquine | 124 |
| ICS_PUSHAsthma | Inhaled steroids | 242 |
| Insulin_Aurum_v2 | Insulin | 188 |
| LABA_PUSH_Asthma | LABA | 166 |
| LAMA_PUSHAsthma | LAMA | 169 |
| LKA_PUSHAsthma | Montelukast | 149 |
| LeflunomideAURUM_Zhaonan | Leflunomide | 76 |
| Levothyroxine_ | Levothyroxine | 252 |
| Lipid_lowering_drugs_Optimal | Lipid lowering drug | 264 |
| Lithium_OPTIMAL | Lithium | 137 |
| MAO_B_Optimal | MAOIs | 87 |
| Meglitinides_Aurum | Meglitinides | 69 |
| Metformin_612_A10BD2 | Metformin | 224 |
| MethotrexateAURUM_Zhaonan | Methotrexate | 147 |
| Mirtazapine | Mirtazapine | 182 |
| Monoamine_Oxidase_Inhibitors_OPTIMAL | MAOIs | 53 |
| NSAIDS_oral_OPTIMAL_final | NSAID | 260 |
| Nasal_steroids_optimal | Nasal steroid | 189 |
| OCS_PUSHAsthma2 | Oral steroids | 192 |
| POP_reg_contraceptive | POP | 157 |
| Paracetamol_OPTIMAL | Paracetamol | 230 |
| Pregabalin_Optimal | Pregabalin | 162 |
| PropranololAnxiety_mumpredict | Propanolol | 160 |
| Rivastigmine | Rivastigmine | 92 |
| SABA_PUSHAsthma | SABA | 234 |
| SGLT2inhibitors_Optimal | SGLT2 inhibitor | 131 |
| SNRIs_OPTIMAL | SNRI | 178 |
| SSRIs_Optimal | SSRI | 254 |
| Statins | Statin | 263 |
| Strong_Opioids_2022_Final | Strong opiates | 197 |
| SulfasalazineAURUM_Zhaonan | Sulfasalazine | 132 |
| Sulphonylureas_Aurum | Sulphonylureas | 200 |
| Systemic_oral_corticosteroids_optimal | Oral steroids | 199 |
| Thiazide_Diuretics_v2 | Thiazides | 249 |
| Thiazolidinediones_Aurum_JC | TZDs | 143 |
| Tricyclic_Antidepressants_final | TCA | 218 |
| UricAcid_LoweringDrugs_optimal | Allopurinol | 177 |
| ValproateMigraine | Sodium valproate | 161 |
| Vitamin_D | Vitamin D supplement | 222 |

*Continued on next page*

Supplementary Table S1 – continued from previous page

| Event category | Plotting label | Token |
|---|---|---|
| Warfarin_ | Warfarin | 203 |
| Weak_Opioids_Final | Weak opiate | 255 |
| all_contraceptive | Contraceptives | 211 |
| asthma_combined_inhalers | Combination inhaler | 190 |
| levodopa_Optimal | Levodopa | 148 |
| phosphodiesterasetype5inhibitor_OPTIMAL | Phosphodiesterase inhibitor | 167 |
| 25_Hydroxyvitamin_D2_level_92 | Vitamin D | 84 |
| 25_Hydroxyvitamin_D3_level_90 | Vitamin D | 86 |
| AST___aspartate_transam_SGOT__46 | AST | 107 |
| AST_serum_level_47 | AST | 154 |
| Albumin___creatinine_ratio_37 | urine ACR | 42 |
| Basophil_count_22 | Basophils | 231 |
| Blood_calcium_level_38 | Calcium | 63 |
| Blood_urea_28 | Urea | 85 |
| Body_mass_index_3 | BMI | 247 |
| Brain_natriuretic_peptide_level_66 | BNP | 51 |
| Calcium_adjusted_level_41 | Calcium | 103 |
| Calculated_LDL_cholesterol_level_103 | LDL | 146 |
| Combined_total_vitamin_D2_and_D3_level_93 | Vitamin D | 66 |
| Corrected_serum_calcium_level_42 | Calcium | 172 |
| Current_smoker_83 | Smoker | 152 |
| Diastolic_blood_pressure_5 | DBP | 261 |
| Eosinophil_count_21 | Eosinophils | 235 |
| Erythrocyte_sedimentation_rate_61 | ESR | 193 |
| Ex_smoker_84 | Ex-smoker | 184 |
| Free_T4_level_76 | T4 | 101 |
| GFR_calculated_abbreviated_MDRD_34 | eGFR | 223 |
| Haematocrit___PCV_16 | Haematocrit | 135 |
| Haematocrit_15 | Haematocrit | 226 |
| Haemoglobin_A1c_level___IFCC_standardised_6 | HbA1c | 202 |
| Haemoglobin_A1c_level_8 | HbA1c | 145 |
| Haemoglobin_estimation_9 | Haemoglobin | 244 |
| HbA1c_level__DCCT_aligned__7 | HbA1c | 165 |
| INR___international_normalised_ratio_81 | INR | 64 |
| International_normalised_ratio_82 | INR | 185 |
| Lymphocyte_count_20 | Lymphocytes | 238 |
| Mean_corpusc_Hb_conc__MCHC__14 | MCHC | 220 |
| Mean_corpusc_haemoglobin_MCH__13 | MCHC | 232 |
| Mean_corpuscular_volume__MCV__11 | MCV | 240 |
| Monocyte_count_23 | Monocytes | 237 |
| N_terminal_pro_brain_natriuretic_peptide_level_67 | BNP | 17 |
| Neutrophil_count_19 | Neutrophils | 239 |
| Never_smoked_tobacco_85 | Never smoker | 217 |
| Non_HDL_cholesterol_level_108 | Non-HDL | 118 |
| O_E___height_1 | Height | 219 |
| O_E___weight_2 | Weight | 251 |
| Plasma_B_natriuretic_peptide_level_69 | BNP | 25 |
| Plasma_C_reactive_protein_60 | CRP | 150 |
| Plasma_HDL_cholesterol_level_101 | HDL | 111 |
| Plasma_LDL_cholesterol_level_104 | LDL | 102 |
| Plasma_N_terminal_pro_B_type_natriuretic_peptid... | BNP | 15 |

*Continued on next page*

Supplementary Table S1 – continued from previous page

| Event category | Plotting label | Token |
|---|---|---|
| Plasma_TSH_level_73 | TSH | 110 |
| Plasma_alanine_aminotransferase_level_44 | ALT | 130 |
| Plasma_albumin_level_52 | Albumin | 128 |
| Plasma_alkaline_phosphatase_level_49 | ALP | 122 |
| Plasma_calcium_level_40 | Calcium | 99 |
| Plasma_cholesterol_HDL_ratio_96 | Cholesterol: HDL ratio | 77 |
| Plasma_corrected_calcium_level_43 | Calcium | 96 |
| Plasma_creatinine_level_32 | Creatinine | 140 |
| Plasma_ferritin_level_62 | Ferritin | 50 |
| Plasma_free_T4_level_77 | T4 | 80 |
| Plasma_gamma_glutamyl_transferase_level_58 | GGT | 108 |
| Plasma_potassium_level_27 | Potassium | 138 |
| Plasma_pro_brain_natriuretic_peptide_level_64 | BNP | 27 |
| Plasma_sodium_level_25 | Sodium | 139 |
| Plasma_total_bilirubin_level_54 | Bilirubin | 125 |
| Plasma_total_cholesterol_level_99 | Total cholesterol | 117 |
| Plasma_triglyceride_level_106 | Triglycerides | 105 |
| Plasma_urea_level_30 | Urea | 109 |
| Platelet_count_12 | Platelets | 243 |
| Red_blood_cell__RBC__count_10 | RBC | 236 |
| Red_blood_cell_distribution_width_17 | RDW | 208 |
| Serum_25_Hydroxy_vitamin_D3_level_88 | Vitamin D | 98 |
| Serum_C_reactive_protein_level_59 | CRP | 176 |
| Serum_HDL_cholesterol_level_100 | HDL | 209 |
| Serum_LDL_cholesterol_level_102 | LDL | 194 |
| Serum_N_terminal_pro_B_type_natriuretic_peptide... | BNP | 61 |
| Serum_T4_level_78 | T4 | 73 |
| Serum_TSH_level_71 | TSH | 214 |
| Serum_alanine_aminotransferase_level_45 | AST | 206 |
| Serum_albumin_51 | Albumin | 229 |
| Serum_alkaline_phosphatase_50 | ALP | 225 |
| Serum_bilirubin_level_53 | Bilirubin | 183 |
| Serum_calcium_39 | Calcium | 186 |
| Serum_cholesterol_97 | Cholesterol | 215 |
| Serum_cholesterol_HDL_ratio_94 | Cholesterol: HDL ratio | 187 |
| Serum_creatinine_31 | Creatinine | 248 |
| Serum_ferritin_63 | Ferritin | 179 |
| Serum_folate_80 | Folic acid | 163 |
| Serum_free_T4_level_75 | T4 | 180 |
| Serum_gamma_glutamyl_transferase_level_57 | GGT | 181 |
| Serum_non_high_density_lipoprotein_cholesterol_... | Non-HDL | 153 |
| Serum_potassium_26 | Potassium | 245 |
| Serum_pro_brain_natriuretic_peptide_level_65 | BNP | 54 |
| Serum_sodium_24 | Sodium | 246 |
| Serum_total_25_hydroxy_vitamin_D_level_87 | Vitamin D | 112 |
| Serum_total_bilirubin_level_56 | Bilirubin | 212 |
| Serum_total_cholesterol_level_98 | Cholesterol | 141 |
| Serum_triglycerides_105 | Triglycerides | 204 |
| Serum_urea_level_29 | Urea | 227 |
| Serum_vitamin_B12_79 | Vitamin B12 | 173 |
| Serum_vitamin_D2_level_89 | Vitamin D | 55 |
| Serum_vitamin_D_86 | Vitamin D | 119 |

*Continued on next page*

Supplementary Table S1 – continued from previous page

| Event category | Plotting label | Token |
|---|---|---|
| Systolic_blood_pressure_4 | SBP | 262 |
| TSH___thyroid_stim_hormone_72 | TSH | 127 |
| TSH_level_74 | TSH | 71 |
| Total_25_hydroxyvitamin_D_level_91 | Vitamin D | 59 |
| Total_alkaline_phosphatase_48 | ALP | 144 |
| Total_bilirubin_55 | Bilirubin | 113 |
| Total_cholesterol_HDL_ratio_95 | Cholesterol: HDL ratio | 170 |
| Total_white_cell_count_18 | WCC | 241 |
| Urine_albumin_creatinine_ratio_35 | Urine ACR | 168 |
| Urine_microalbumin_creatinine_ratio_36 | Urine ACR | 47 |
| eGFR_using_creatinine_CKD_EPI_per_1_73_square_m... | eGFR | 159 |

**Supplementary Table S2: Event counts.** Number of clinical records used for pre-training, collected from over $26.5$ million patients across $1478$ practices in England. Counts are extracted from the SQLite database, and the count of observed values indicates the number available before outlier removal.

| Event type | Number of events | Token count | Count of observed values |
|---|---|---|---|
| Diagnoses | 74 | 51,003,640 | n/a |
| Medications | 81 | 3,970,984,804 | n/a |
| Investigations | 108 | 3,533,426,831 | 3,364,437,065 |
| **Total** | 263 | 7.6 Billion | 3.4 Billion |

**Supplementary Table S3: Summary statistics of diagnoses.** Summary statistics of raw diagnosis data extracted from CPRD by DExtER. These are calculated before any pre-processing has been performed.

| Event category | Count |
|---|---|
| ADDISONS_DISEASE | 6691 |
| ADDISON_DISEASE | 11794 |
| AF | 731332 |
| ALCOHOLMISUSE_V2 | 1125212 |
| ALLCANCER_NOHAEM_NOBCC | 1496973 |
| ALLERGICRHINITISCONJ | 3291165 |
| ALL_DEMENTIA | 528602 |
| ANXIETY | 3560978 |
| ANY_DEAFNESS_HEARING_LOSS_V2 | 2282766 |
| AORTICANEURYSM_V2 | 101134 |
| ASTHMA_PUSHASTHMA | 4175115 |
| ATOPICECZEMA | 4369082 |
| AUTISM | 156860 |
| BIPOLAR | 108852 |
| BRONCHIECTASIS | 112618 |
| CHRONICFATIGUESYNDROMEMM_V2 | 82799 |
| CHRONIC_LIVER_DISEASE_ALCOHOL | 63405 |
| CKDSTAGE3TO5 | 1088754 |
| COPD | 751320 |
| CROHNS_DISEASE | 81250 |
| CYSTICFIBROSIS | 7053 |
| DEATH | 1629100 |

*Continued on next page*

Supplementary Table S3 – continued from previous page

| Event category | Count |
|---|---|
| DEPRESSION | 4109336 |
| DOWNSSYNDROME | 17006 |
| EATINGDISORDERS | 191873 |
| ENDOMETRIOSIS_ADENOMYOSIS_V2 | 209157 |
| EPILEPSY | 377341 |
| FIBROMYALGIA | 153213 |
| GOUT | 632089 |
| HAEMOCHROMATOSIS_V2 | 18631 |
| HF_V3 | 524982 |
| HIVAIDS | 41951 |
| HYPERTENSION | 3934473 |
| HYPERTHYROIDISM_V2 | 217964 |
| HYPOTHYROIDISM_DRAFT_V1 | 932079 |
| IHDINCLUDINGMI_OPTIMALV2 | 1162843 |
| ILD_SH | 58104 |
| ISCHAEMICSTROKE_V2 | 178437 |
| LEUKAEMIA_PREVALENCEV2 | 54438 |
| LYMPHOMA_PREVALENCE_V2 | 80511 |
| MENIERESDISEASE | 73688 |
| MINFARCTION | 477556 |
| MS | 53204 |
| NAFLD_V2 | 158688 |
| OSA | 234938 |
| OSTEOARTHRITIS | 2653242 |
| OSTEOPOROSIS | 628118 |
| OTHER_CHRONIC_LIVER_DISEASE_OPTIMAL | 289252 |
| PAD_STRICT | 254632 |
| PARKINSONS | 91718 |
| PERIPHERAL_NEUROPATHY | 840638 |
| PERNICIOUSANAEMIA | 73331 |
| PLASMACELL_NEOPLASM_V2 | 20301 |
| PMRANDGCA | 195907 |
| POLYCYSTIC_OVARIAN_SYNDROME_PCOS_V2 | 332297 |
| PREVALENT_IBS_V2 | 1210229 |
| PSORIASIS | 743897 |
| PSORIATICARTHRITIS2021 | 51273 |
| PTSDDIAGNOSIS | 126269 |
| PVD_V3 | 185904 |
| RHEUMATOIDARTHRITIS | 197253 |
| SCHIZOPHRENIAMM_V2 | 125141 |
| SICKLE_CELL_DISEASE_V2 | 11159 |
| SJOGRENSSYNDROME | 23326 |
| STROKEUNSPECIFIED_V2 | 446046 |
| STROKE_HAEMRGIC | 83609 |
| SUBSTANCEMISUSE | 502552 |
| SYSTEMIC_LUPUS_ERYTHEMATOSUS | 26820 |
| SYSTEMIC_SCLEROSIS | 8772 |
| TYPE1DM | 145143 |
| TYPE2DIABETES | 1404325 |
| ULCERATIVE_COLITIS | 120361 |
| VALVULARDISEASES_V2 | 401061 |
| VISUAL_IMPAIRMENT | 155707 |

**Supplementary Table S4: Summary statistics of medications.** Summary statistics of raw medication data extracted from CPRD by DExtER. These are calculated before any pre-processing has been performed.

| Event category | Count |
|---|---|
| ACE_Inhibitors_D2T | 191462111 |
| AMD_DH_Donepezil | 5635153 |
| AMD_DH_Memantine | 2309471 |
| ARBs_Luyuan | 72329278 |
| Acarbose_AURUM | 507434 |
| AldosteroneAntagonist_D2T | 11182704 |
| AllHIVdrugs_HIV | 82262 |
| All_AntiArrhythmics_D2T | 29502352 |
| All_Antiplatelets | 162940629 |
| All_Diuretics_D2T | 176070847 |
| All_Diuretics_ExclLactones_D2T | 176070847 |
| AlphaBlocker | 51635379 |
| Amantadine | 479791 |
| Amitriptyline_optimal | 46415710 |
| Anticonvulsants_OPTIMAL | 90774779 |
| Antihistamines_second_generation_H1_antagonists | 47308115 |
| Antipsychotics_OPTIMAL | 31658166 |
| Anxiolytics_mumpredict | 32874923 |
| Aspirin_OPTIMAL | 131327634 |
| Benzodiazepines | 55608827 |
| BetaBlockers_OPTIMAL | 79374522 |
| Bisphosphonates_osteoporosis | 30294675 |
| COCP_reg_contraception | 34316525 |
| COMT_Optimal | 886259 |
| CalciumChannelBlck_D2T | 108721302 |
| Calcium_supplements_Optimal | 57542223 |
| Carbamazepine_Optimal | 12812897 |
| DPP4inhibitors_OPTIMAL | 12000147 |
| Dementia | 9738544 |
| Digoxin_ | 17497910 |
| Direct_Acting_Oral_Anticoagulants_DOAC | 14815747 |
| Fibrate_FenofibrateOnly | 2305308 |
| First_gen_H1_antihistamines | 16181607 |
| GLP_1Aurum | 2676211 |
| Gabapentin_Oral_OPTIMAL | 17508445 |
| Galantamine_NEW | 830429 |
| HydroxychloroquineAURUM_Zhaonan | 3312310 |
| ICS_PUSHAsthma | 94873269 |
| Insulin_Aurum_v2 | 28641496 |
| LABA_PUSH_Asthma | 14958767 |
| LAMA_PUSHAsthma | 15239995 |
| LKA_PUSHAsthma | 8537907 |
| LeflunomideAURUM_Zhaonan | 586799 |
| Levothyroxine_ | 117844926 |
| Lipid_lowering_drugs_Optimal | 263876709 |
| Lithium_OPTIMAL | 4395155 |
| MAO_B_Optimal | 829174 |
| Meglitinides_Aurum | 486644 |
| Metformin_612_A10BD2 | 74162080 |
| MethotrexateAURUM_Zhaonan | 7884622 |

*Continued on next page*

Supplementary Table S4 – continued from previous page

| Event category | Count |
|---|---|
| Mirtazapine | 23524446 |
| Monoamine_Oxidase_Inhibitors_OPTIMAL | 236966 |
| NSAIDS_oral_OPTIMAL_final | 219183349 |
| Nasal_steroids_optimal | 29010523 |
| OCS_PUSHAsthma2 | 29568929 |
| POP_reg_contraceptive | 12186107 |
| Paracetamol_OPTIMAL | 85242874 |
| Pregabalin_Optimal | 14497547 |
| PropranololAnxiety_mumpredict | 12924874 |
| Rivastigmine | 963471 |
| SABA_PUSHAsthma | 91182585 |
| SGLT2inhibitors_Optimal | 4059940 |
| SNRIs_OPTIMAL | 21379065 |
| SSRIs_Optimal | 132417423 |
| Statins | 249911634 |
| Strong_Opioids_2022_Final | 31728471 |
| SulfasalazineAURUM_Zhaonan | 4084855 |
| Sulphonylureas_Aurum | 34289555 |
| Systemic_oral_corticosteroids_optimal | 33516650 |
| Thiazide_Diuretics_v2 | 105228802 |
| Thiazolidinediones_Aurum_JC | 6191596 |
| Tricyclic_Antidepressants_final | 68893019 |
| UricAcid_LoweringDrugs_optimal | 20559558 |
| ValproateMigraine | 13594697 |
| Vitamin_D | 72657845 |
| Warfarin_ | 37353354 |
| Weak_Opioids_Final | 140622354 |
| all_contraceptive | 53655141 |
| asthma_combined_inhalers | 29469516 |
| levodopa_Optimal | 8483033 |
| phosphodiesterasetype5inhibitor_OPTIMAL | 15057609 |

**Supplementary Table S5: Summary statistics of investigations.** Summary statistics of raw investigation data extracted from CPRD by DExtER. These are calculated before any pre-processing has been performed.

| Event category | Mean | Min | Max | Count | # Missing |
|---|---|---|---|---|---|
| 25_Hydroxyvitamin_D2_level_92 | 3.91 | 0 | 686 | 782791 | 89321 |
| 25_Hydroxyvitamin_D3_level_90 | 47.1 | 0 | 952 | 809104 | 27986 |
| AST___aspartate_transam_SGOT__46 | 26.6 | 0 | 1.5e+04 | 1738489 | 57876 |
| AST_serum_level_47 | 27.3 | -5.00 | 2.1e+04 | 10837982 | 352631 |
| Albumin___creatinine_ratio_37 | 10.7 | -1.00 | 1.3e+04 | 180911 | 102491 |
| Basophil_count_22 | 0.0501 | -0.100 | 1.1e+05 | 86869779 | 1227239 |
| Blood_calcium_level_38 | 2.35 | 0 | 440 | 415717 | 30253 |
| Blood_urea_28 | 6.51 | 0 | 1.3e+03 | 785766 | 113905 |
| Body_mass_index_3 | 293 | -3.3e+04 | 2.1e+09 | 99868822 | 2109510 |
| Brain_natriuretic_peptide_level_66 | 417 | 0 | 5e+05 | 229202 | 69884 |
| Calcium_adjusted_level_41 | 2.37 | 0 | 279 | 1464226 | 6315 |
| Calculated_LDL_cholesterol_level_103 | 2.82 | -8.40 | 240 | 6967780 | 248034 |
| Combined_total_vitamin_D2_and_D3_level_93 | 52.4 | 0 | 1.2e+03 | 452984 | 28416 |

*Continued on next page*

Supplementary Table S5 – continued from previous page

| Event category | Mean | Min | Max | Count | # Missing |
|---|---|---|---|---|---|
| Corrected_serum_calcium_level_42 | 3.89 | -2.58 | 1.2e+07 | 16683796 | 419151 |
| Current_smoker_83 | 69.8 | -2e+03 | 1.7e+07 | 10592787 | 8083175 |
| Diastolic_blood_pressure_5 | 3.8e+03 | -120 | 9.1e+11 | 241026671 | 301816 |
| Eosinophil_count_21 | 0.234 | -0.600 | 4.4e+05 | 92193433 | 584466 |
| Erythrocyte_sedimentation_rate_61 | 18.4 | -20.0 | 1.3e+07 | 29892785 | 5233008 |
| Ex_smoker_84 | 410 | -2e+03 | 2e+08 | 24891632 | 22903077 |
| Free_T4_level_76 | 2.9e+05 | -134 | 9.8e+10 | 1405341 | 150687 |
| GFR_calculated_abbreviated_MDRD_34 | 72.0 | -90.0 | 1.8e+07 | 73709249 | 7876443 |
| Haematocrit___PCV_16 | 5.57 | -0.465 | 9.4e+03 | 4346108 | 32185 |
| Haematocrit_15 | 3.78 | 0 | 2.1e+05 | 76582325 | 287307 |
| Haemoglobin_A1c_level___IFCC_standard... | 1.9e+05 | -79.0 | 6e+12 | 35091904 | 2549714 |
| Haemoglobin_A1c_level_8 | 13.5 | 0 | 1e+06 | 6516701 | 4237674 |
| Haemoglobin_estimation_9 | 79.6 | -112 | 1.4e+07 | 97391693 | 858067 |
| HbA1c_level__DCCT_aligned__7 | 11.7 | -41.0 | 3.3e+07 | 14948615 | 2828072 |
| INR___international_normalised_ratio_81 | 2.78 | 0 | 3.1e+04 | 426165 | 16990 |
| International_normalised_ratio_82 | 23.9 | -34.0 | 3e+07 | 27966697 | 3393266 |
| Lymphocyte_count_20 | 365 | -35.0 | 3.7e+09 | 92896401 | 526313 |
| Mean_corpusc_Hb_conc__MCHC__14 | 186 | -295 | 3.5e+05 | 70867102 | 242847 |
| Mean_corpusc_haemoglobin_MCH__13 | 82.8 | -28.9 | 3.6e+07 | 90498376 | 277562 |
| Mean_corpuscular_volume__MCV__11 | 2.1e+04 | -100 | 2e+12 | 94723195 | 313276 |
| Monocyte_count_23 | 81.9 | -4.00 | 3.7e+09 | 92489278 | 510098 |
| N_terminal_pro_brain_natriuretic_peptide_level_67 | 760 | 0 | 7.5e+04 | 57470 | 4069 |
| Neutrophil_count_19 | 85.1 | -67.0 | 3.7e+09 | 93489124 | 637721 |
| Never_smoked_tobacco_85 | 5.09 | -25.0 | 2e+03 | 60751822 | 59213176 |
| Non_HDL_cholesterol_level_108 | 3.45 | -1.60 | 3.4e+03 | 2408310 | 14410 |
| O_E___height_1 | 664 | -1.1e+05 | 2e+10 | 69089436 | 336366 |
| O_E___weight_2 | 5.8e+04 | -1.2e+04 | 4e+12 | 117401338 | 589408 |
| Plasma_B_natriuretic_peptide_level_69 | 215 | 0 | 5.7e+05 | 82488 | 3711 |
| Plasma_C_reactive_protein_60 | 13.7 | -3.8e+03 | 1.3e+07 | 9155551 | 1754490 |
| Plasma_HDL_cholesterol_level_101 | 1.8e+03 | 0 | 3.7e+09 | 2173456 | 61682 |
| Plasma_LDL_cholesterol_level_104 | 2.84 | 0 | 502 | 1434461 | 184941 |
| Plasma_N_terminal_pro_B_type_natriuretic_peptid... | 832 | 0.200 | 1e+05 | 53534 | 1229 |
| Plasma_TSH_level_73 | 2.59 | -7.80 | 2.5e+04 | 2049380 | 32179 |
| Plasma_alanine_aminotransferase_level_44 | 26.8 | 0 | 1.1e+04 | 4012790 | 23486 |
| Plasma_albumin_level_52 | 40.8 | 0 | 4.4e+04 | 3719377 | 11968 |
| Plasma_alkaline_phosphatase_level_49 | 128 | -24.0 | 2e+04 | 3290802 | 9406 |
| Plasma_calcium_level_40 | 2.38 | 0 | 378 | 1294409 | 7535 |
| Plasma_cholesterol_HDL_ratio_96 | 3.79 | 0 | 999 | 587483 | 11890 |
| Plasma_corrected_calcium_level_43 | 2.35 | 0 | 248 | 1168224 | 268132 |
| Plasma_creatinine_level_32 | 915 | 0 | 3.7e+09 | 4513054 | 23789 |
| Plasma_ferritin_level_62 | 84.2 | 0 | 2.5e+04 | 220380 | 52482 |
| Plasma_free_T4_level_77 | 5.7e+05 | -77.2 | 2.5e+10 | 724108 | 42413 |
| Plasma_gamma_glutamyl_transferase_level_58 | 53.6 | 0 | 1.2e+04 | 1762750 | 13310 |
| Plasma_potassium_level_27 | 849 | 0 | 3.7e+09 | 4445152 | 42312 |
| Plasma_pro_brain_natriuretic_peptide_level_64 | 550 | 0 | 6.7e+04 | 83343 | 13214 |
| Plasma_sodium_level_25 | 980 | 0 | 3.7e+09 | 4458632 | 33102 |
| Plasma_total_bilirubin_level_54 | 11.0 | 0 | 1.1e+04 | 3443114 | 13631 |
| Plasma_total_cholesterol_level_99 | 1.6e+03 | 0 | 3.7e+09 | 2331589 | 28898 |
| Plasma_triglyceride_level_106 | 2.4e+03 | 0 | 3.7e+09 | 1573268 | 24409 |
| Plasma_urea_level_30 | 5.96 | 0 | 808 | 1991274 | 22825 |
| Platelet_count_12 | 2.5e+04 | -560 | 3.7e+09 | 95904116 | 521873 |
| Red_blood_cell__RBC__count_10 | 6.1e+04 | -4.46 | 3.7e+09 | 92326200 | 470623 |

*Continued on next page*

Supplementary Table S5 – continued from previous page

| Event category | Mean | Min | Max | Count | # Missing |
|---|---|---|---|---|---|
| Red_blood_cell_distribution_width_17 | 14.0 | 0 | 2.6e+05 | 49751137 | 3364137 |
| Serum_25_Hydroxy_vitamin_D3_level_88 | 50.0 | 0 | 2e+04 | 1211141 | 363513 |
| Serum_C_reactive_protein_level_59 | 23.8 | -115 | 2.1e+08 | 19234068 | 2893243 |
| Serum_HDL_cholesterol_level_100 | 1.51 | -39.0 | 1.8e+06 | 50953183 | 936362 |
| Serum_LDL_cholesterol_level_102 | 387 | -1e+06 | 3.7e+09 | 30073152 | 981676 |
| Serum_N_terminal_pro_B_type_natriuretic_peptide... | 786 | 0 | 1.2e+05 | 383790 | 48134 |
| Serum_T4_level_78 | 5.2e+06 | -25.0 | 1.5e+11 | 524777 | 73789 |
| Serum_TSH_level_71 | 15.4 | -21.4 | 2e+07 | 55655662 | 3197511 |
| Serum_alanine_aminotransferase_level_45 | 27.1 | -43.0 | 2.1e+05 | 46896239 | 777411 |
| Serum_albumin_51 | 791 | -45.0 | 9e+09 | 83105055 | 349161 |
| Serum_alkaline_phosphatase_50 | 89.3 | -165 | 1.5e+07 | 76052242 | 342655 |
| Serum_bilirubin_level_53 | 16.2 | -1.9e+05 | 1.2e+08 | 24090337 | 147279 |
| Serum_calcium_39 | 2.64 | -2.12 | 7e+06 | 28049902 | 1120505 |
| Serum_cholesterol_97 | 74.7 | -75.0 | 3.7e+09 | 57393841 | 2256817 |
| Serum_cholesterol_HDL_ratio_94 | 137 | -3.40 | 3.7e+09 | 28586515 | 637258 |
| Serum_creatinine_31 | 535 | -4.2e+04 | 3.7e+09 | 102915170 | 1314960 |
| Serum_ferritin_63 | 120 | -2.8e+06 | 1.9e+08 | 21412371 | 2581319 |
| Serum_folate_80 | 8.60 | -20.0 | 2.4e+04 | 14643748 | 1398479 |
| Serum_free_T4_level_75 | 5.5e+05 | 0 | 1e+11 | 22579062 | 600152 |
| Serum_gamma_glutamyl_transferase_level_57 | 57.4 | -462 | 7e+04 | 22592131 | 788041 |
| Serum_non_high_density_lipoprotein_cholesterol_... | 3.40 | -2.50 | 7.3e+03 | 10648096 | 27356 |
| Serum_potassium_26 | 43.2 | -2.2e+03 | 3.7e+09 | 99036754 | 1540838 |
| Serum_pro_brain_natriuretic_peptide_level_65 | 701 | 0 | 7.7e+05 | 243565 | 47507 |
| Serum_sodium_24 | 141 | -140 | 6.9e+07 | 99409745 | 592495 |
| Serum_total_25_hydroxy_vitamin_D_level_87 | 53.2 | 0 | 921 | 2178432 | 296091 |
| Serum_total_bilirubin_level_56 | 32.0 | -2.00 | 4.3e+07 | 54098383 | 344930 |
| Serum_total_cholesterol_level_98 | 5.17 | -7.60 | 1e+06 | 5185017 | 84358 |
| Serum_triglycerides_105 | 445 | -376 | 3.7e+09 | 42415036 | 434218 |
| Serum_urea_level_29 | 5.8e+03 | -24.1 | 2.4e+10 | 78670253 | 393889 |
| Serum_vitamin_B12_79 | 715 | -5.8e+08 | 3.7e+09 | 16797696 | 1665415 |
| Serum_vitamin_D2_level_89 | 14.8 | -5.00 | 728 | 249092 | 44373 |
| Serum_vitamin_D_86 | 53.7 | -39.7 | 3e+06 | 2478806 | 737039 |
| Systolic_blood_pressure_4 | 601 | -200 | 1.1e+11 | 241541988 | 318809 |
| TSH___thyroid_stim_hormone_72 | 25.6 | -50.0 | 8.1e+06 | 3568579 | 505048 |
| TSH_level_74 | 4.04 | -0.500 | 9.8e+03 | 507085 | 234183 |
| Total_25_hydroxyvitamin_D_level_91 | 48.0 | 0 | 1.2e+03 | 333324 | 49869 |
| Total_alkaline_phosphatase_48 | 84.5 | 0 | 9e+03 | 6461505 | 38583 |
| Total_bilirubin_55 | 147 | -7.00 | 1.8e+07 | 2193031 | 51422 |
| Total_cholesterol_HDL_ratio_95 | 965 | -3.10 | 3.7e+09 | 15760772 | 271673 |
| Total_white_cell_count_18 | 250 | -14.7 | 3.7e+09 | 94827537 | 648527 |
| Urine_albumin_creatinine_ratio_35 | 996 | -14.0 | 1e+10 | 15107807 | 4858601 |
| Urine_microalbumin_creatinine_ratio_36 | 10.8 | -5.80 | 3.2e+04 | 201318 | 107309 |
| eGFR_using_creatinine_CKD_EPI_... | 74.3 | -90.0 | 8.7e+04 | 12869245 | 967096 |

**Supplementary Table S6: Pre-processing bounds.** Outlier and standardisation bounds. Values beyond these limits are treated as outliers and marked as missing by SurvivEHR. Remaining values are standardised between these bounds, centered at zero.

| Event category | $F_L$ | $F_U$ |
|---|---|---|
| 25_Hydroxyvitamin_D2_level_92 | -4.67 | 10.9 |
| 25_Hydroxyvitamin_D3_level_90 | -36.8 | 122 |
| AST___aspartate_transam_SGOT__46 | 0.100 | 40000 |
| AST_serum_level_47 | 0.100 | 40000 |
| Albumin___creatinine_ratio_37 | -4.32 | 8.83 |
| Basophil_count_22 | -0.0938 | 10.0 |
| Blood_calcium_level_38 | 0.100 | 10.0 |
| Blood_urea_28 | 0.100 | 11.1 |
| Body_mass_index_3 | 5.00 | 130 |
| Brain_natriuretic_peptide_level_66 | -245 | 483 |
| Calcium_adjusted_level_41 | 0.100 | 10.0 |
| Calculated_LDL_cholesterol_level_103 | 0 | 30.0 |
| Combined_total_vitamin_D2_and_D3_level_93 | 0.100 | 127 |
| Corrected_serum_calcium_level_42 | 0.100 | 10.0 |
| Current_smoker_83 | 0 | 80.0 |
| Diastolic_blood_pressure_5 | 20.0 | 400 |
| Eosinophil_count_21 | -0.185 | 10.0 |
| Erythrocyte_sedimentation_rate_61 | 0 | 49.7 |
| Ex_smoker_84 | 0 | 80.0 |
| Free_T4_level_76 | 0.100 | 1000 |
| GFR_calculated_abbreviated_MDRD_34 | 0.100 | 300 |
| Haematocrit___PCV_16 | 0.100 | 1.00 |
| Haematocrit_15 | 0 | 1.00 |
| Haemoglobin_A1c_level___IFCC_standardised_6 | 5.00 | 250 |
| Haemoglobin_A1c_level_8 | 5.00 | 250 |
| Haemoglobin_estimation_9 | 10.0 | 300 |
| HbA1c_level__DCCT_aligned__7 | 5.00 | 250 |
| INR___international_normalised_ratio_81 | 0.625 | 100 |
| International_normalised_ratio_82 | 0.425 | 100 |
| Lymphocyte_count_20 | 0.100 | 200 |
| Mean_corpusc_Hb_conc__MCHC__14 | 10.0 | 500 |
| Mean_corpusc_haemoglobin_MCH__13 | 1.00 | 100 |
| Mean_corpuscular_volume__MCV__11 | 10.0 | 250 |
| Monocyte_count_23 | -0.0153 | 20.0 |
| N_terminal_pro_brain_natriuretic_peptide_level_67 | -609 | 1182 |
| Neutrophil_count_19 | 0 | 50.0 |
| Never_smoked_tobacco_85 | 0 | 0 |
| Non_HDL_cholesterol_level_108 | 0 | 6.41 |
| O_E___height_1 | 20.0 | 272 |
| O_E___weight_2 | 0.250 | 650 |
| Plasma_B_natriuretic_peptide_level_69 | -179 | 368 |
| Plasma_C_reactive_protein_60 | -11.2 | 22.9 |
| Plasma_HDL_cholesterol_level_101 | 0.343 | 3.90 |
| Plasma_LDL_cholesterol_level_104 | 0 | 30.0 |
| Plasma_N_terminal_pro_B_type_natriuretic_peptid... | -634 | 1243 |
| Plasma_TSH_level_73 | -1.34 | 500 |
| Plasma_alanine_aminotransferase_level_44 | 0.100 | 40000 |
| Plasma_albumin_level_52 | 2.00 | 180 |
| Plasma_alkaline_phosphatase_level_49 | 1.00 | 281 |

*Continued on next page*

Supplementary Table S6 – continued from previous page

| Event category | $F_L$ | $F_U$ |
|---|---|---|
| Plasma_calcium_level_40 | 0.100 | 10.0 |
| Plasma_cholesterol_HDL_ratio_96 | 1.00 | 250 |
| Plasma_corrected_calcium_level_43 | 0.100 | 1.00 |
| Plasma_creatinine_level_32 | 1.00 | 140 |
| Plasma_ferritin_level_62 | -90.1 | 500000 |
| Plasma_free_T4_level_77 | 0.100 | 1000 |
| Plasma_gamma_glutamyl_transferase_level_58 | -26.3 | 90.7 |
| Plasma_potassium_level_27 | 0.100 | 10.0 |
| Plasma_pro_brain_natriuretic_peptide_level_64 | -347 | 670 |
| Plasma_sodium_level_25 | 90.0 | 200 |
| Plasma_total_bilirubin_level_54 | -0.841 | 900 |
| Plasma_total_cholesterol_level_99 | 1.64 | 50.0 |
| Plasma_triglyceride_level_106 | -0.488 | 3.46 |
| Plasma_urea_level_30 | 0.100 | 10.1 |
| Platelet_count_12 | 0 | 10000000 |
| Red_blood_cell__RBC__count_10 | 0 | 500 |
| Red_blood_cell_distribution_width_17 | 10.4 | 100 |
| Serum_25_Hydroxy_vitamin_D3_level_88 | 0.100 | 124 |
| Serum_C_reactive_protein_level_59 | -8.03 | 17.4 |
| Serum_HDL_cholesterol_level_100 | 0 | 3.90 |
| Serum_LDL_cholesterol_level_102 | 0 | 30.0 |
| Serum_N_terminal_pro_B_type_natriuretic_peptide... | -616 | 1198 |
| Serum_T4_level_78 | 0.100 | 1000 |
| Serum_TSH_level_71 | -1.16 | 500 |
| Serum_alanine_aminotransferase_level_45 | 0.100 | 40000 |
| Serum_albumin_51 | 2.00 | 180 |
| Serum_alkaline_phosphatase_50 | 1.00 | 20000 |
| Serum_bilirubin_level_53 | -0.918 | 900 |
| Serum_calcium_39 | 0.100 | 10.0 |
| Serum_cholesterol_97 | 1.76 | 50.0 |
| Serum_cholesterol_HDL_ratio_94 | 1.00 | 250 |
| Serum_creatinine_31 | 1.00 | 10000 |
| Serum_ferritin_63 | -113 | 500000 |
| Serum_folate_80 | -4.58 | 20.5 |
| Serum_free_T4_level_75 | 0.100 | 10000 |
| Serum_gamma_glutamyl_transferase_level_57 | -29.0 | 94.8 |
| Serum_non_high_density_lipoprotein_cholesterol_... | 0 | 3.90 |
| Serum_potassium_26 | 0.100 | 10.0 |
| Serum_pro_brain_natriuretic_peptide_level_65 | -500 | 931 |
| Serum_sodium_24 | 90.0 | 200 |
| Serum_total_25_hydroxy_vitamin_D_level_87 | -19.2 | 124 |
| Serum_total_bilirubin_level_56 | -0.908 | 900 |
| Serum_total_cholesterol_level_98 | 1.57 | 50.0 |
| Serum_triglycerides_105 | 0 | 100 |
| Serum_urea_level_29 | 0.100 | 10.2 |
| Serum_vitamin_B12_79 | 5.00 | 20000 |
| Serum_vitamin_D2_level_89 | -7.75 | 20.6 |
| Serum_vitamin_D_86 | 0.100 | 1000 |
| Systolic_blood_pressure_4 | 30.0 | 400 |
| TSH___thyroid_stim_hormone_72 | -1.53 | 500 |
| TSH_level_74 | -1.96 | 500 |
| Total_25_hydroxyvitamin_D_level_91 | 0.100 | 1000 |

*Continued on next page*

Supplementary Table S6 – continued from previous page

| Event category | $F_L$ | $F_U$ |
|---|---|---|
| Total_alkaline_phosphatase_48 | 0.100 | 139 |
| Total_bilirubin_55 | -2.32 | 900 |
| Total_cholesterol_HDL_ratio_95 | 1.00 | 250 |
| Total_white_cell_count_18 | 0.100 | 5000 |
| Urine_albumin_creatinine_ratio_35 | -3.78 | 300 |
| Urine_microalbumin_creatinine_ratio_36 | -4.12 | 8.25 |
| eGFR_using_creatinine_CKD_EPI_... | 22.7 | 300 |

#### Figures

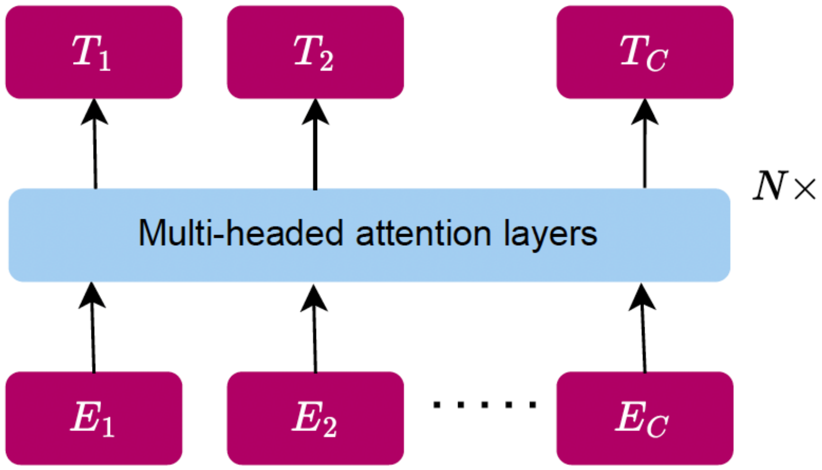

**Supplementary Figure S1: Decoder architecture.** SurvivEHR uses the Generative Pre-trained Transformer (GPT) decoder architecture to learn representations of a patient's prior health records. At any point in time all historic events $\mathbf{E}_1, \ldots \mathbf{E}_c$ can be processed to obtain a vector embedding that summarises the patient's history, $\mathbf{T}_c$.

| Component | Hyperparameter | Pre-training | Fine-tuning |
|---|---|---|---|
| Architecture | Layers | 6 | 6 |
| | Attention heads | 6 | 6 |
| | Hidden size | 384 | 384 |
| | Max sequence length | 256 | 512 |
| Context window | Repeated measurements | All | Last unique |
| | Global diagnoses | False | Append to context |
| Optimization | Batch size | 64 | 512 |
| | Epochs | 10 | 20 |
| | Early stopping | False | True |
| | Optimizer | AdamW | AdamW |
| | Backbone learning rate | $3 \times 10^{-4}$ | $5 \times 10^{-5}$ |
| | Head learning rate | $3 \times 10^{-4}$ | $5 \times 10^{-4}$ |
| Scheduler | Linear warmup steps | 10,000 | 0 |
| | Schedule | Cosine annealing | Reduce on plateau |
| | Warm restarts | True | False |
| | Warmup steps | 10,000 | - |
| | LR decay | 0.8 | - |

**Supplementary Table S7:** Model hyperparameters used during pre-training and fine-tuning.

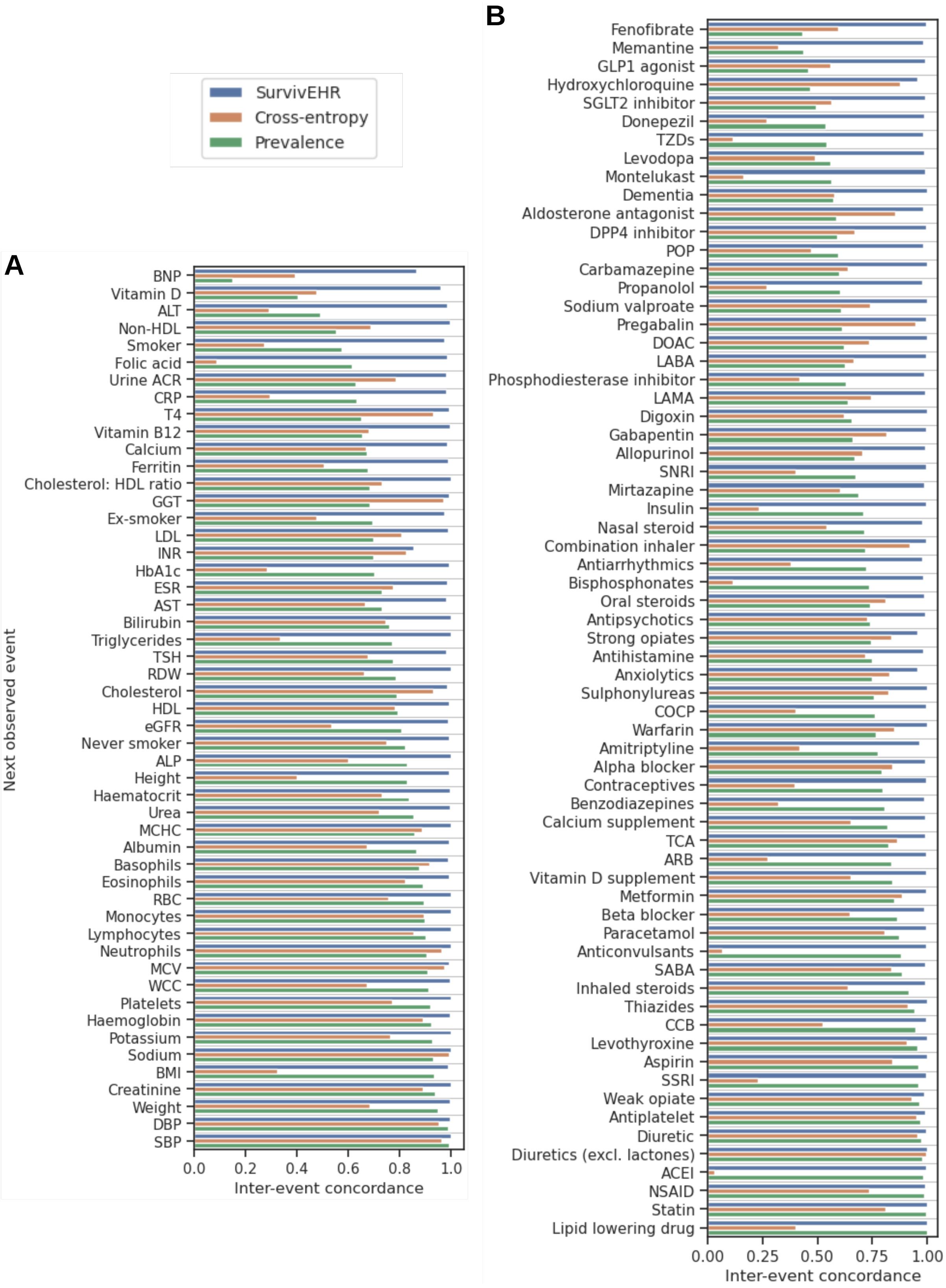

**Supplementary Figure S2: Inter-event concordance.** The inter-event concordance metric for each **A** investigation, and **B** medication. This quantifies the model's ability to correctly rank the risk of each diagnosis being the next-event a patient will experience across their lifetime. Some tokens are missing due to random sampling.

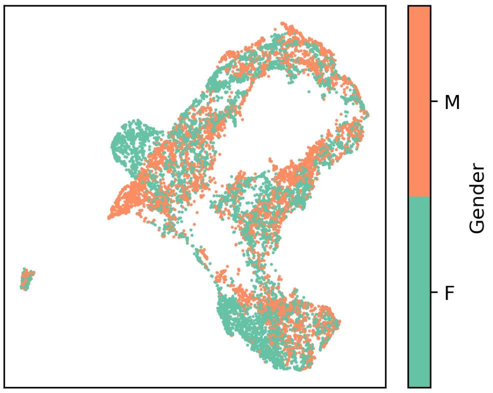

**(a)** Sex.

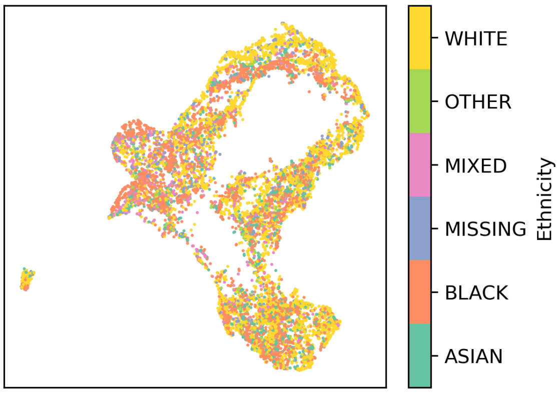

**(b)** Ethnicity.

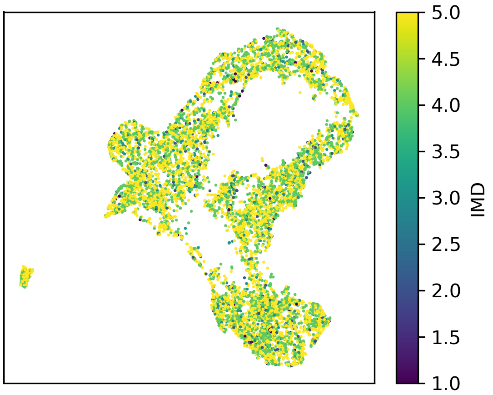

**(c)** Index of multiple deprivation.

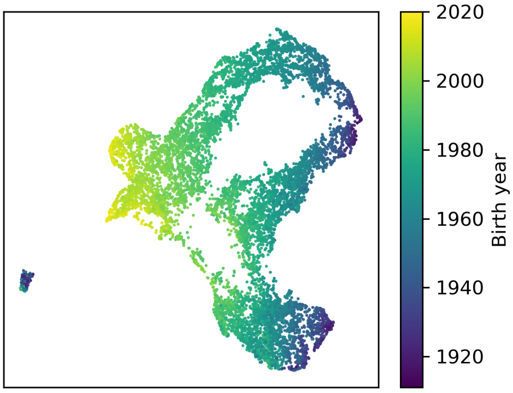

**(d)** Year of birth.

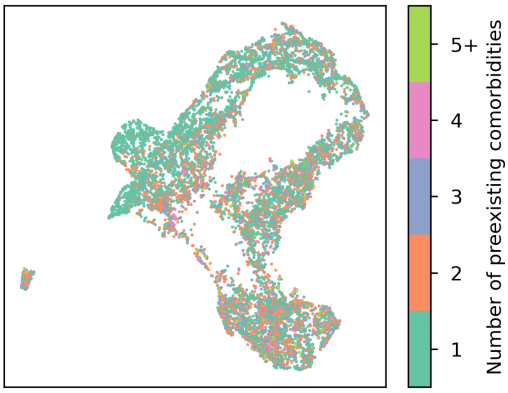

**(e)** Number of pre-existing conditions.

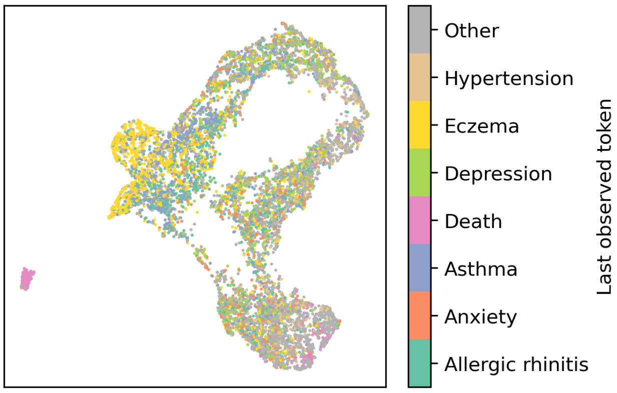

**(f)** Last observed diagnosis.

**Supplementary Figure S3: Learnt representations.** UMAP representations $\mathbf{T}_c$ of patient health records in the held-out pre-training cohort. We include histories up to and including each diagnosis a patient experiences.

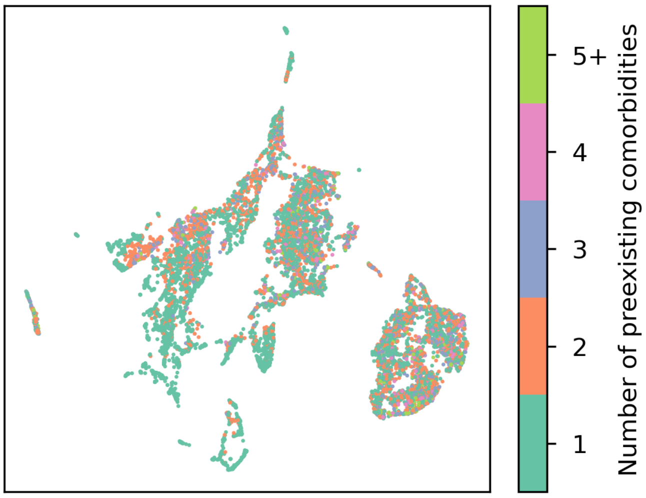

**(a)** Number of pre-existing conditions.

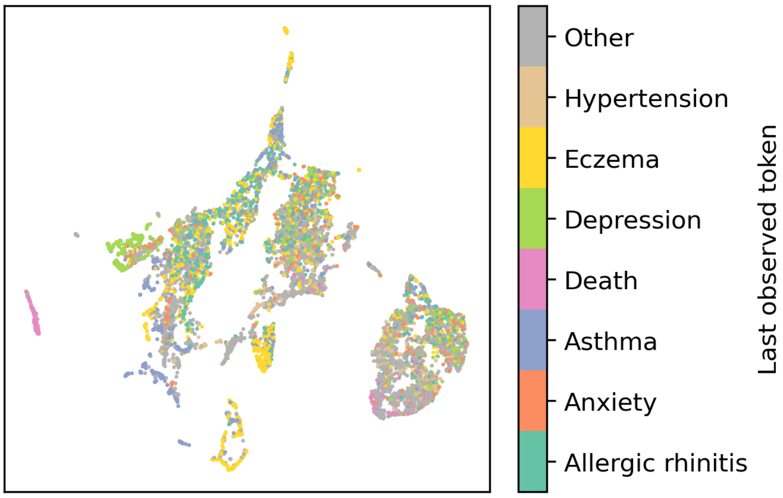

**(b)** Last observed diagnosis.

**Supplementary Figure S4: Learnt representations with baseline covariates masked.** UMAP representations $\mathbf{T}_c$ of patient health records in the held-out pre-training cohort with baseline covariates $\mathbf{e}^{\text{static}}$ removed. We include histories up to and including each diagnosis a patient experiences.

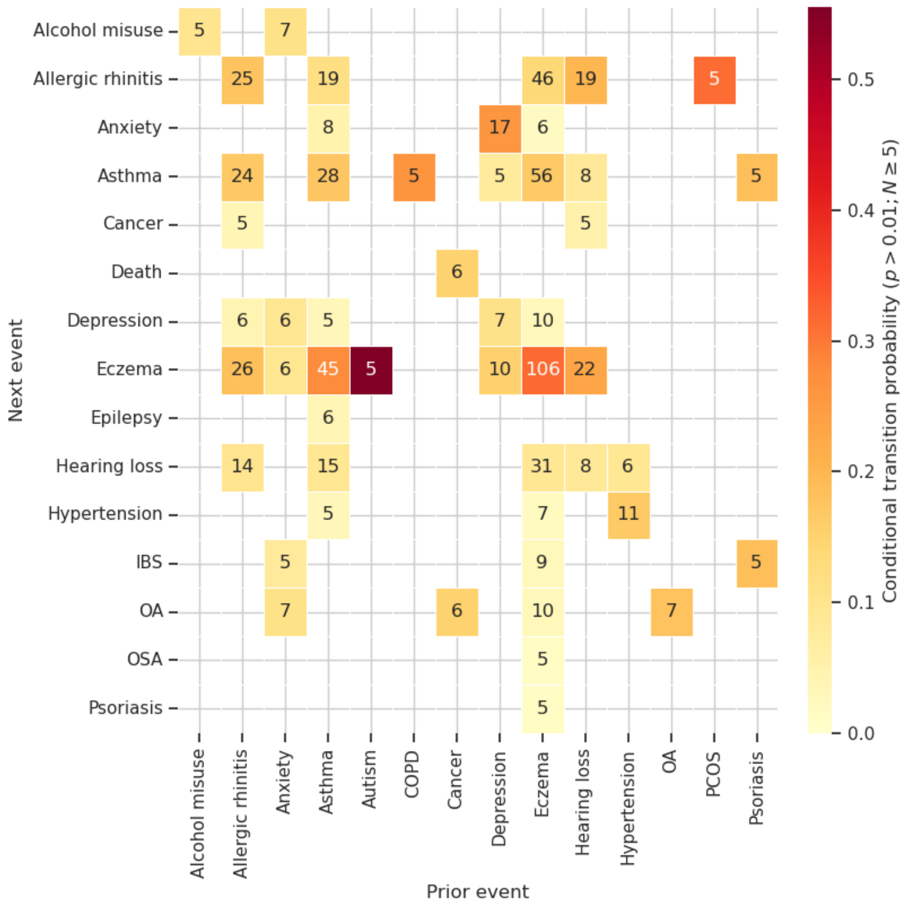

**Supplementary Figure S5: Forecasting known diagnosis to diagnosis associations in the pre-training cohort.** Given unseen patient histories from the pre-training cohort, we predict the next 3 events a patient may experience. From these we plot the transitional probability matrices for a new diagnosis following a previous diagnosis. Transitions with fewer than $5$ occurrences and probability below $0.01$ are removed for clarity.

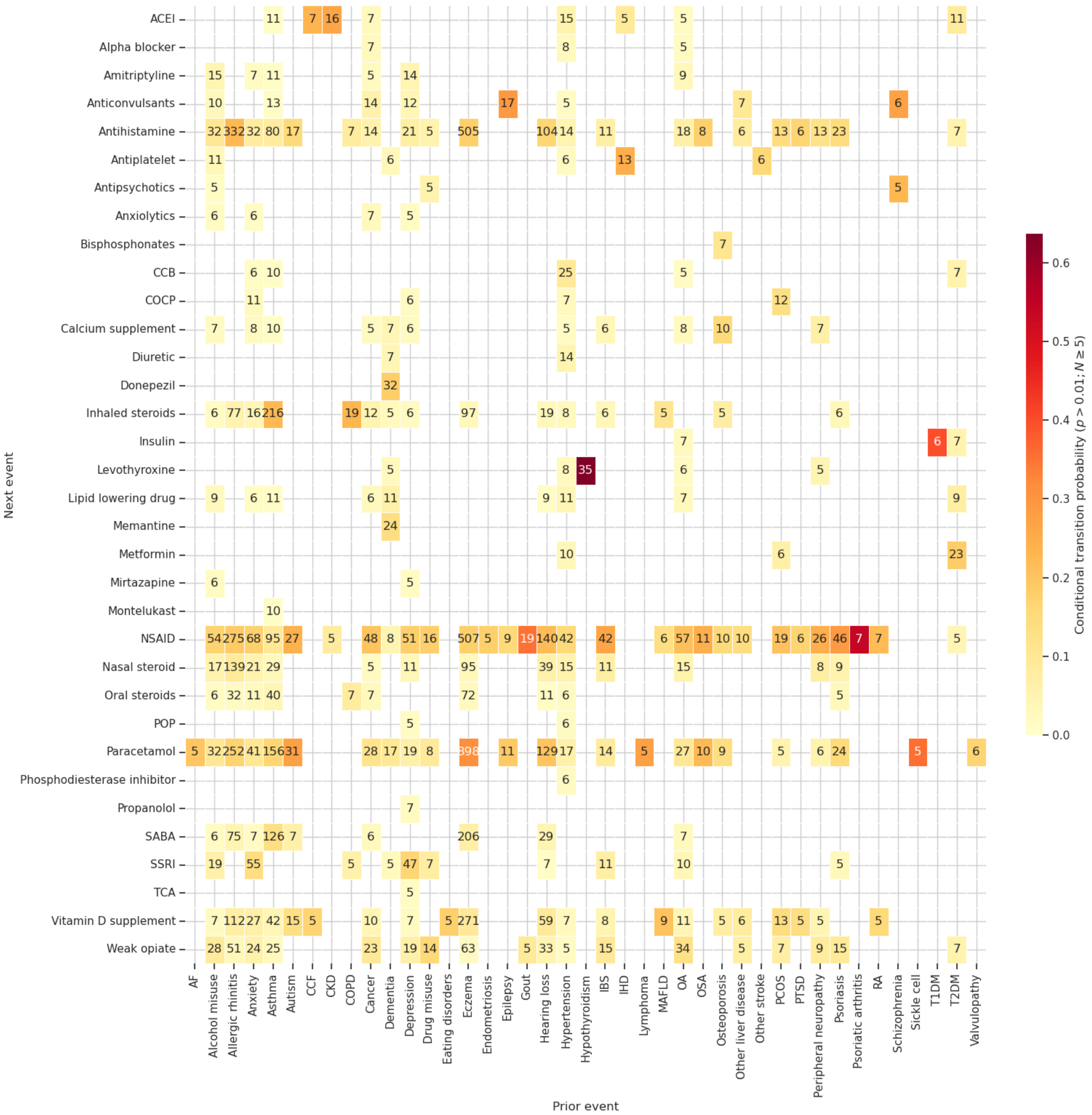

**Supplementary Figure S6: Forecasting known diagnosis to medication associations in the pre-training cohort.** Given unseen patient histories from the pre-training cohort, we predict the next 3 events a patient may experience. From these we plot the transitional probability matrices for a new medication following a previous diagnosis. Transitions with fewer than $5$ occurrences and probability below $0.01$ are removed for clarity.

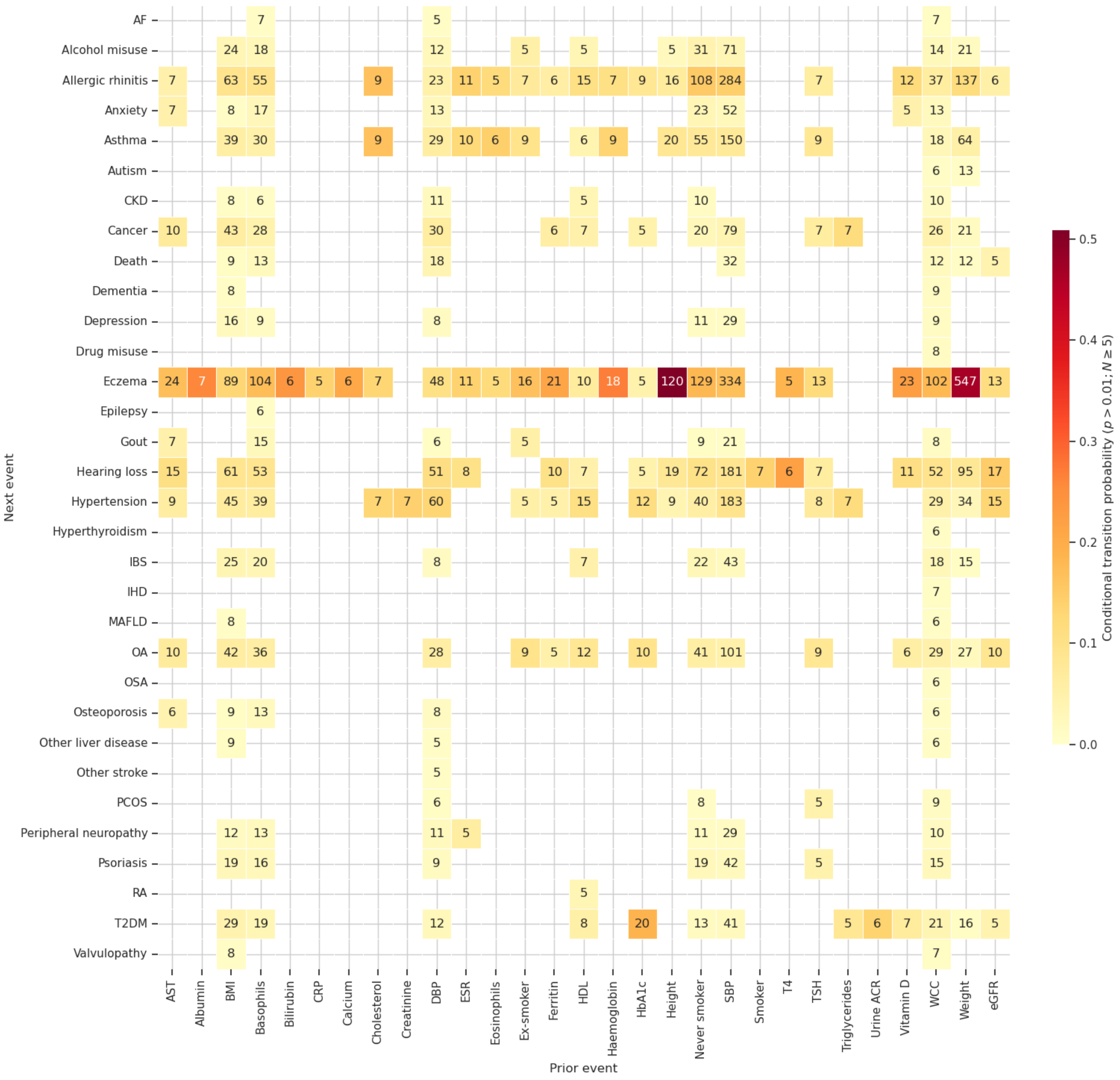

**Supplementary Figure S7: Forecasting known investigation to diagnosis associations in the pre-training cohort.** Given unseen patient histories from the pre-training cohort, we predict the next 3 events a patient may experience. From these we plot the transitional probability matrices for a new diagnosis following a previous investigation. Transitions with fewer than $5$ occurrences and probability below $0.01$ are removed for clarity.
